# Evaluating the effectiveness of rapid SARS-CoV-2 genome sequencing in supporting infection control teams: the COG-UK hospital-onset COVID-19 infection study

**DOI:** 10.1101/2022.02.10.22270799

**Authors:** Oliver Stirrup, James Blackstone, Fiona Mapp, Alyson MacNeil, Monica Panca, Alison Holmes, Nicholas Machin, Gee Yen Shin, Tabitha Mahungu, Kordo Saeed, Tranprit Saluja, Yusri Taha, Nikunj Mahida, Cassie Pope, Anu Chawla, Maria-Teresa Cutino-Moguel, Asif Tamuri, Rachel Williams, Alistair Darby, David Robertson, Flavia Flaviani, Eleni Nastouli, Samuel Robson, Darren Smith, Matthew Loose, Kenneth Laing, Irene Monahan, Beatrix Kele, Sam Haldenby, Ryan George, Matthew Bashton, Adam Witney, Matthew Byott, Francesc Coll, Michael Chapman, Sharon Peacock, COG-UK HOCI Investigators, The COVID-19 Genomics UK (COG-UK) consortium, Joseph Hughes, Gaia Nebbia, David G Partridge, Matthew Parker, James Price, Christine Peters, Sunando Roy, Luke B Snell, Thushan I de Silva, Emma Thomson, Paul Flowers, Andrew Copas, Judith Breuer

**Affiliations:** Institute for Global Health, UCL, London, UK; Comprehensive Clinical Trials Unit, UCL, London, UK; Imperial College Healthcare NHS Trust, London, UK; Manchester University NHS Foundation Trust, Manchester, UK; University College London Hospitals NHS Foundation Trust, London, UK; Royal Free NHS Foundation Trust, London, UK; University Hospital Southampton NHS Foundation Trust, Southampton, UK, and Clinical and Experimental Sciences, School of Medicine, University of Southampton; Sandwell and West Birmingham NHS Trust, UK; Departments of Virology and Infectious Diseases, Newcastle upon Tyne Hospitals NHS Foundation Trust, Newcastle, UK; Nottingham University Hospitals NHS Trust, Nottingham, UK; St George’s University Hospitals NHS Foundation Trust, London, UK; Liverpool University Hospitals NHS Foundation Trust, Liverpool, UK; Barts Health NHS Trust, London, UK; Research Computing, UCL, London, UK; Department of Genetics & Genomic Medicine, UCL Great Ormond Street Institute of Child Health, UCL, London, UK; Centre for Genomic Research, University of Liverpool, Liverpool, UK; MRC-University of Glasgow Centre for Virus Research, Glasgow, UK; Guy’s and St Thomas’ Hospital NHS Foundation Trust, London, UK; Centre for Enzyme Innovation and School of Pharmacy and Biomedical Science,, University of Portsmouth, Portsmouth, UK; Department of Applied Sciences, Northumbria University, Newcastle, UK; School of Life Sciences, University of Nottingham, Nottingham, UK; Institute for Infection and Immunity, St George’s University of London, London, UK; The Hub for Biotechnology in the Built Environment, Department of Applied Sciences, Northumbria University, Newcastle, UK; Department of Infection Biology, Faculty of Infectious and Tropical Diseases, London School of Hygiene & Tropical Medicine, London, UK; Health Data Research UK Cambridge Hub, Cambridge UK; Department of Medicine, University of Cambridge, Cambridge, UK; Sheffield Teaching Hospitals NHS Foundation Trust, Sheffield, UK; Sheffield Bioinformatics Core, The University of Sheffield, Sheffield, UK; NHS Greater Glasgow and Clyde, Glasgow, UK; Department of Infection, Immunity and Inflammation, UCL Great Ormond Street Institute of Child Health, UCL, London, UK; Department of Infection, Immunity and Cardiovascular Disease, Medical School, The University of Sheffield, Sheffield, UK; School of Psychological Sciences and Health, University of Strathclyde, Glasgow, UK

**Keywords:** COVID-19, viral genomics, hospital-acquired infection, healthcare-associated infection, infection prevention and control, molecular epidemiology, SARS-CoV-2

## Abstract

**Introduction:** Viral sequencing of SARS-CoV-2 has been used for outbreak investigation, but there is limited evidence supporting routine use for infection prevention and control (IPC) within hospital settings.

**Methods:** We conducted a prospective non-randomised trial of sequencing at 14 acute UK hospital trusts. Sites each had a 4-week baseline data-collection period, followed by intervention periods comprising 8 weeks of ‘rapid’ (<48h) and 4 weeks of ‘longer-turnaround’ (5-10 day) sequencing using a sequence reporting tool (SRT). Data were collected on all hospital onset COVID-19 infections (HOCIs; detected ≥48h from admission). The impact of the sequencing intervention on IPC knowledge and actions, and on incidence of probable/definite hospital-acquired infections (HAIs) was evaluated.

**Results:** A total of 2170 HOCI cases were recorded from October 2020-April 2021, with sequence reports returned for 650/1320 (49.2%) during intervention phases. We did not detect a statistically significant change in weekly incidence of HAIs in longer-turnaround (IRR 1.60, 95%CI 0.85-3.01; *P=*0.14) or rapid (0.85, 0.48-1.50; *P=*0.54) intervention phases compared to baseline phase. However, IPC practice was changed in 7.8% and 7.4% of all HOCI cases in rapid and longer-turnaround phases, respectively, and 17.2% and 11.6% of cases where the report was returned. In a per-protocol sensitivity analysis there was an impact on IPC actions in 20.7% of HOCI cases when the SRT report was returned within 5 days.

**Conclusion:** While we did not demonstrate a direct impact of sequencing on the incidence of nosocomial transmission, our results suggest that sequencing can inform IPC response to HOCIs, particularly when returned within 5 days.

## Introduction

Viral sequencing has played an important role in developing our understanding of the emergence and evolution of the SARS-CoV-2 pandemic^[1]^. Sequencing technologies can now be used for local outbreak investigation in near real-time, and this was implemented by some research centres for evaluation of nosocomial transmission from the early stages of the pandemic^[2]^. It has been demonstrated that sequencing can provide additional information on outbreak characteristics and transmission routes in comparison to traditional epidemiological investigation alone^[2-4]^. However, limited data are available on the feasibility of routine use of sequencing for infection prevention and control (IPC), or on its direct impact on IPC actions and nosocomial transmission rates.

Throughout the pandemic, nosocomial transmission of SARS-CoV-2 has been a major concern^[5]^, with hospital-acquired infections (HAIs) accounting for more than 5% of lab-confirmed cases from March-August 2020 in the UK^[6]^ and representing 11% of COVID-19 cases within hospitals in this period^[7]^. HAIs also frequently occur within a very vulnerable population with high levels of mortality^[6, 8, 9]^. There is therefore an unmet need to develop interventions that can reduce the occurrence of nosocomial transmission. The aims of this study were to determine the effectiveness of SARS-CoV-2 sequencing in informing acute IPC actions and reducing the incidence of HAIs when used prospectively in routine practice, and to record the impact of sequencing reports on the actions of IPC teams.

## Methods

We conducted a prospective multiphase non-randomised trial to evaluate the implementation and impact of SARS-CoV-2 sequencing for IPC within 14 acute NHS hospital groups in the UK. All sites were linked to a COG-UK sequencing hub, 13 were university hospitals and one a district general hospital. We implemented a bespoke sequence reporting tool (SRT) intervention, developed and previously evaluated for this study^[10]^, and assessed the importance of turnaround time for sequencing and reporting. The study included integral health economic and qualitative process evaluation^[11]^.

The study design comprised a planned 4-week baseline data-collection period, followed by intervention periods defined by the time from diagnostic sampling to return of sequence data to IPC teams, comprising 8 weeks of ‘rapid’ (<48 hours) turnaround sequencing and 4 weeks of ‘longer’ (5-10 days) turnaround sequencing for each site. Target turnaround time was 48h from diagnostic sampling to return of the SRT report during the ‘rapid’ sequencing phase, and 5-10 days for the ‘longer-turnaround’ phase. Eight sites implemented rapid’ followed by ‘longer-turnaround’ phases with five doing the opposite. One site did not implement longer-turnaround sequencing because they considered it a reduction in their standard practice. Data were recorded in all phases for all patients meeting the definition of: a hospital-onset COVID-19 infection (HOCI), i.e. first confirmed test for SARS-CoV-2 >48 hours after admission and without suspicion of COVID-19 at time of admission. During the intervention phases, and for at least 3 weeks prior to any intervention period to enable linkage to recent cases, participating sites aimed to sequence all SARS-CoV-2 cases including both HOCI and non-HOCI cases.

The SRT aimed to integrate sequence and patient data to produce concise and immediately interpretable feedback about cases to IPC teams via a one-page report. Sites were also able to apply other methods (e.g. phylogenetics) to the sequence data generated, where this was part of their usual practice.

Data collection on patient characteristics and on implementation and impact of the intervention was conducted using a central study-specific database. Ethical approval for the study was granted by NHS HRA (REC 20/EE/0118), and the study was prospectively registered (ClinicalTrials.gov Identifier: NCT04405934).

The primary outcomes of the study as defined in the protocol^[12]^ were: (1) incidence of IPC-defined SARS-CoV-2 HAIs per week per 100 currently admitted non-COVID-19 inpatients, and (2) for each HOCI, identification of linkage to individuals within an outbreak of SARS-CoV-2 nosocomial transmission using sequencing data as interpreted through the SRT that was not identified by pre-sequencing IPC evaluation during intervention phases.

Secondary outcomes were: (1) incidence of IPC-defined SARS-CoV-2 hospital outbreaks per week per 100 non-COVID-19 inpatients, (2) for each HOCI, any change to IPC actions following receipt of SRT report during intervention phases, (3) any recommended change to IPC actions (regardless of whether changes were implemented). The proportion of HOCI cases for which IPC reported the SRT report to be ‘useful’ was added as a further outcome.

To support standardisation across sites, ‘IPC-defined SARS-CoV-2 HAIs’ were considered to be all HOCIs with ≥8 days from admission to symptom onset (if known) or sample date (i.e. UK Health Security Agency definition of a probable/definite HAI^[13]^).

An IPC-defined SARS-CoV-2 hospital outbreak was defined as at least two HOCI cases on the same ward, with at least one having ≥8 days from admission to symptom onset or sample date. Outbreak events were considered to be concluded once there was a period of 28 days prior to observation of another HOCI^[13]^.

Further details of outcome definitions are given in the Supplementary Appendix.

### Statistical analysis

We used three approaches: intention-to-treat analysis to assess the overall impact of sequencing on IPC activity and the incidence of HAIs, per-protocol site-based analysis on a subset of high performance sites, and pooled analysis to describe how turnaround time was related to impact on IPC irrespective of study phase. Inclusion of sites in the per-protocol analysis was based on the proportion of sequence reports returned and speed of return in the rapid phase. Thresholds to define this group were determined following review of the data but before analysis of outcomes.

Incidence outcomes were analysed using mixed effects negative binomial models. Data for the first week of each intervention period, or in the first week of return to intervention following a break, were considered transition periods and not considered as direct evidence regarding the intervention effect. Analysis was conducted with calendar time divided into ‘study weeks’ running Monday-Sunday. Models were adjusted for calendar time, the proportion of current inpatients that were SARS-CoV-2 positive, as well as local community SARS-CoV-2 incidence for each study site, using 5-knot restricted cubic splines ^[14]^. Number of inpatients not positive for SARS-CoV-2 was considered an exposure variable (defining ‘person-time’ at risk of nosocomial infection). Differences between study phases were evaluated using adjusted incidence rate ratios.

The primary outcome of identification of SARS-CoV-2 nosocomial transmission using sequencing data and secondary outcomes relating to changes to IPC actions were analysed using mixed effects logistic regression models. Marginal proportions from fitted models were reported for rapid- and longer-turnaround intervention phases, and differences in outcomes between these phases were evaluated. If the SRT report was not returned this was interpreted as a ‘failure’, i.e. no change to IPC action; however, we also present percentages for these outcomes restricted to HOCIs where the SRT report was returned.

For both incidence and ‘per HOCI’ outcomes, we accounted for the structure of the data with hierarchical exchangeable normally distributed random effects for each study site, and for each study phase within each study site. Analyses were conducted using Stata V16, with figures generated using the *ggplot2* package for R V4.0.

## Results

A total of 2170 HOCIs were recorded for the study between 15 October 2020 and 26 April 2021. These cases had median age of 76.7 (IQR 64.4-85.6) years, and 80% had at least one clinically significant comorbidity (Table 1).

**Table 1:** Demographic and baseline characteristics of the participants by study phase

| Characteristic at screening | Study phase |  |  | Total |
| --- | --- | --- | --- | --- |
|  | Baseline | Longer-turnaround | Rapid |  |
| *N* HOCl cases | 850 | 373 | 947 | 2170 |
| *N* HOCl cases per site, median (range); *N* sites | 36 (1-207); 14 | 19 (0-86); 13 | 30.5 (4-297); 14 | 103.5 (40-451); 14 |
| HAI classification, *n* (%) |  |  |  |  |
| Indeterminate (3-7 days) | 362 (42.6) | 166 (44.5) | 371 (39.2) | 899 (41.4) |
| Probable (8-14 days) | 236 (27.8) | 121 (32.4) | 270 (28.5) | 627 (28.9) |
| Definite (>14 days) | 252 (29.6) | 86 (23.1) | 306 (32.3) | 644 (29.7) |
| Age (years), median (IQR, range) | 77.5 (65.4-85.6, 0.4-100.5) | 77.6 (64.6-86.7, 0.7-100.7) | 76.4 (62.6-85.5, 0.6-103.5) | 76.7 (64.4-85.6, 0.4-103.5) |
| Age ≥70 years, *n/N* (%) | 589/850 (69.3) | 240/373 (64.3) | 598/947 (63.1) | 1427/2170 (65.8) |
| Sex at birth: female, *n/N* (%) | 457/850 (53.8) | 177/372 (47.6) | 460/947 (48.6) | 1094/2169 (50.4) |
| Ethnicity, *n* (%) |  |  |  |  |
| White | 668 (78.6) | 275 (73.7) | 732 (77.3) | 1675 (77.2) |
| Mixed ethnicity | 9 (1.1) | 6 (1.6) | 8 (0.8) | 23 (1.1) |
| Asian | 46 (5.4) | 26 (7.0) | 34 (3.6) | 106 (4.9) |
| Black Caribbean or African | 36 (4.2) | 18 (4.8) | 46 (4.9) | 100 (4.6) |
| Other | 6 (0.7) | 1 (0.3) | 4 (0.4) | 11 (0.5) |
| Unknown | 85 (10.0) | 47 (12.6) | 123 (13.0) | 255 (11.8) |
| Symptomatic at time of sampling, *n/N* (%) | 167/739 (22.6) | 58/322 (18.0) | 106/659 (16.1) | 331/1720 (19.2) |
| Significant comorbidity present, *n/N* (%) | 650/776 (83.8) | 260/323 (80.5) | 574/757 (75.8) | 1484/1856 (80.0) |
| Pregnant, *n/N* (%) | 6/451 (1.3) | 1/177 (0.6) | 4/445 (0.9) | 11/1073 (1.0) |
| Hosp. admission route, *n* (%) |  |  |  |  |
| Emergency department | 605 (71.2) | 258 (69.2) | 549 (58.0) | 1412 (65.1) |
| Hospital transfer | 59 (6.9) | 21 (5.6) | 51 (5.4) | 131 (6.0) |
| Care home | 3 (0.4) | 0 (0) | 0 (0) | 3 (0.1) |
| GP referral | 38 (4.5) | 15 (4.0) | 76 (8.0) | 129 (5.9) |
| Outpatient clinic ref. | 27 (3.2) | 20 (5.4) | 30 (3.2) | 77 (3.5) |
| Other | 42 (4.9) | 9 (2.4) | 48 (5.1) | 99 (4.6) |
| Unknown | 76 (8.9) | 50 (13.4) | 193 (20.4) | 319 (14.7) |

HAI, hospital-acquired infection; HOCl, hospital onset COVID-19 infection; Hosp., hospital.

All 14 sites completed baseline and rapid sequencing intervention phases (Figure S1). Thirteen sites completed the longer-turnaround sequencing intervention phase. 49.2% (650/1320) SRT reports for HOCIs were returned in the intervention phases, with only 9.3% (123/1320) returned within the target timeframes (Table 2). This figure was greater in the longer-turnaround phase at 21.2% (79/373) than in the rapid phase (4.6%; 44/947). The median turnaround time from diagnostic sampling for reports returned was 5 days in the rapid phase and 13 days in the longer-turnaround phase.

**Table 2:** Per HOCI implementation and outcome summary by study intervention phase, overall and within the 7/14 sites included in the ‘per protocol’ sensitivity analysis

|  | All study sites |  |  | Sensitivity analysis |  |
| --- | --- | --- | --- | --- | --- |
|  | Study phase |  | Total | Study phase |  |
|  | Longer-turnaround | Rapid |  | Longer-turnaround | Rapid |
| *N* HOCl cases | 373 | 947 | 1320 | 143 | 533 |
| **Implementation** |  |  |  |  |  |
| Sequence returned within expected timeline, *n* (%)* | 229 (61.4) | 377 (39.8) | 606 (45.9) | 81 (56.6) | 204 (38.3) |
| Sequence returned within study period, *n* (%)* | 277 (74.3) | 596 (62.9) | 873 (66.1) | 98 (68.5) | 347 (65.1) |
| SRT report returned within target timeline (10d for longer-turnaround, 2d for rapid), *n* (%) | 79 (21.2) | 44 (4.6) | 123 (9.3) | 35 (24.5) | 44 (8.3) |
| SRT report returned within study period, *n* (%) | 215 (57.6) | 435 (45.9) | 650 (49.2) | 92 (64.3) | 317 (59.5) |
| Time from sample to report return (days), median (IQR, range) [*n*] | 13 (9-15, 0-36) [215] | 5 (3-11, 2-84) [430] | 9 (4-14, 0-84) [645] | 13 (9-17, 6-29) [92] | 4 (3-6, 2-64) [312] |
| **Sequencing results** |  |  |  |  |  |
| SRT suggestive patient acquired infection post-admission, *n/N* (%) | 196/212 (92.5) | 384/423 (90.8) | 580/635 (91.3) | 85/92 (92.4) | 287/311 (92.3) |
| SRT suggestive patient is part of ward outbreak, *n/N* (%) | 151/212 (71.2) | 260/423 (61.5) | 411/635 (64.7) | 65/92 (70.7) | 202/311 (65.0) |
| Linkage identified not suspected at initial IPC investigation: |  |  |  |  |  |
| All HOCl in phase *n/N* (%†, 95% CI) | 24/348 (6.8, 1.7-11.8) | 46/915 (6.7, 2.0-11.3) | 70/1263 (5.5) | 11/139 (7.9, 3.4-12.4) | 39/512 (7.6, 5.3-9.9) |
| When SRT returned *n/N* (%) | 24/190 (12.6) | 46/403 (11.4) | 70/593 (11.8) | 11/88 (12.5) | 39/296 (13.2) |
| SRT excluded IPC-identified hospital outbreak, *n/N* (%) | 14/213 (6.6) | 27/428 (6.3) | 41/641 (6.4) | 9/92 (9.8) | 25/310 (8.1) |
| **Impact on IPC** |  |  |  |  |  |
| SRT changed IPC practice: |  |  |  |  |  |
| All HOCl in phase *n/N* (%†, 95% CI) | 25/373 (7.4, 1.1-13.6) | 74/941 (7.8, 2.4-13.2) | 99/1314 (7.5) | 1/143 (0.7, 0.0-2.1) | 52/527 (9.9, 7.3-12.4) |
| When SRT returned *n/N* (%) | 25/215 (11.6) | 74/429 (17.2) | 99/644 (15.4) | 1/92 (1.1) | 52/311 (16.7) |
| SRT changed IPC practice for ward, *n/N* (%) | 13/215 (6.0) | 31/429 (7.2) | 44/644 (6.8) | 0/92 (0.0) | 28/311 (9.0) |
| SRT used in IPC decisions beyond ward, *n/N* (%) | 12/215 (5.6) | 45/428 (10.5) | 57/643 (8.9) | 1/92 (1.1) | 27/310 (8.7) |
| IPC team reported SRT to be useful, *n/N* (%) |  |  |  |  |  |
| Yes | 107/215 (49.8) | 303/428 (70.8) | 410/643 (63.8) | 25/92 (27.2) | 245/310 (79.0) |
| No | 67/215 (31.2) | 71/428 (16.6) | 138/643 (21.5) | 50/92 (54.3) | 57/310 (18.4) |
| Unsure | 41/215 (19.1) | 54/428 (12.6) | 95/643 (14.8) | 17/92 (18.5) | 8/310 (2.6) |
| **HCW absence on ward** |  |  |  |  |  |
| Prop. HCWs on sick leave due to COVID-19, median (IQR, range) [*n*] | 0.09 (0.00-0.15, 0.00-0.30) [49] | 0.13 (0.07-0.29, 0.00-1.00) [162] | 0.13 (0.04-0.27, 0.00-1.00) [321]‡ | 0.09 (0.00-0.15, 0.00-0.30) [49] | 0.13 (0.08-0.29, 0.00-1.00) [143] |

HCW, healthcare worker; IPC, infection prevention and control; IQR, interquartile range; IRR, incidence rate ratio; Prop., proportion; SRT, sequence reporting tool. \*As recorded by site, not based on recorded date or availability on central CLIMB server. †Estimated marginal value from mixed effects model, not raw %, evaluated on intention-to-treat basis with lack of SRT report classified as 'no'. ‡Includes data for baseline phase: 0.13 (0.00-0.30, 0.00-0.88) [110].

Ordering the sites by proportion of cases with sequencing results returned and median turnaround time during the rapid phase (Figure 1) identified no obvious clustering of highest versus lowest performing sites. We therefore also carried out a ‘per protocol’ sensitivity analysis on the seven highest performing sites; these sites returned ≥40% of SRTs within a median time from diagnostic sample of ≤8 days within their rapid phase.

**Figure 1.**
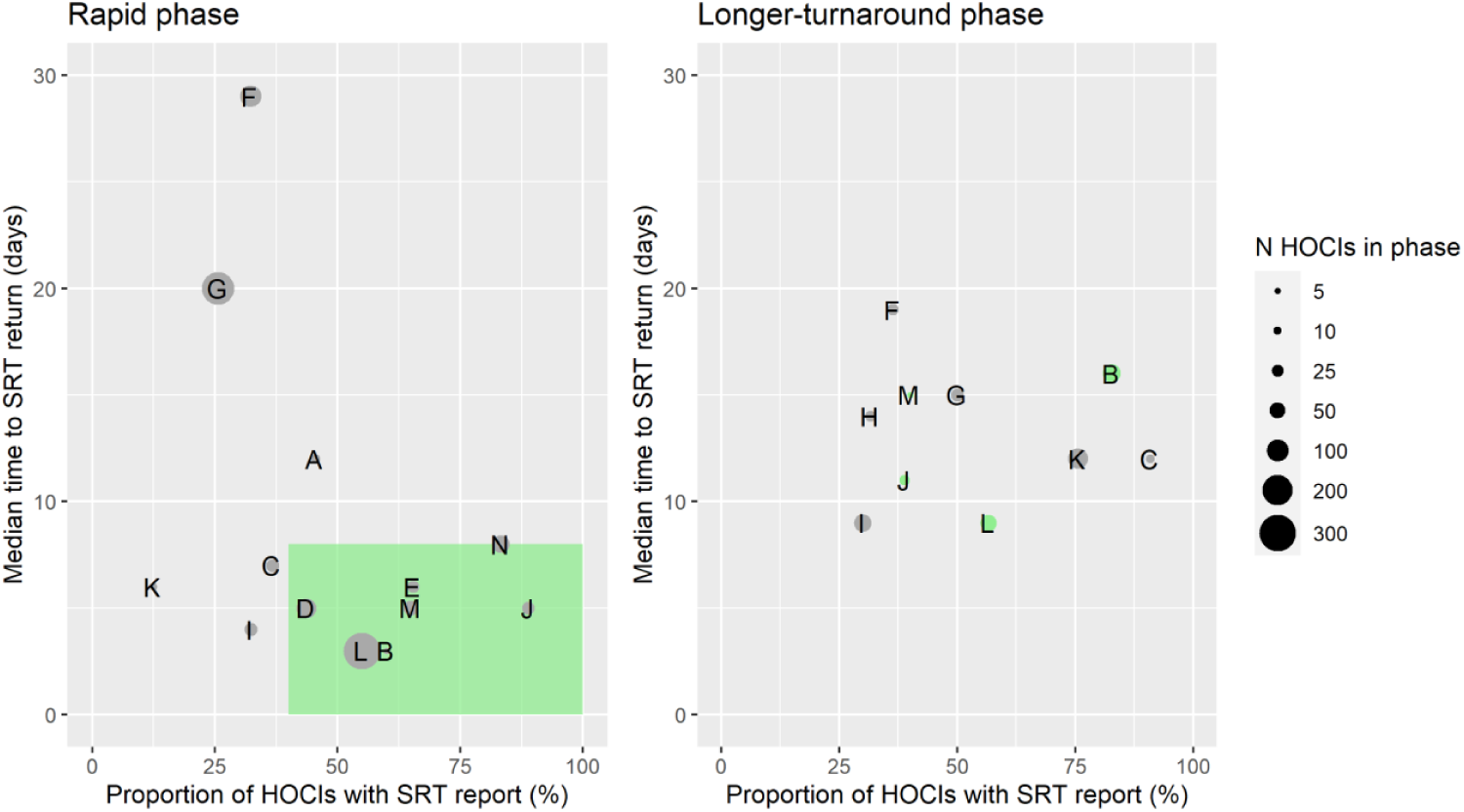
Plots of the median turnaround time against the percentage of HOCI cases with SRT reports returned for the rapid (left panel) and longer-turnaround (right panel) sequencing phases across the 14 study sites. The size of each circle plotted indicates the number of HOCI cases observed within each phase for each site, with letter labels corresponding to study site. The criteria for inclusion in our sensitivity analysis are displayed as the green rectangle in the rapid phase plot, and sites on the longer-turnaround phase plot are color-coded by their inclusion. In the rapid phase, SRT reports were returned for 0/4 HOCI cases recorded for Site H. Site N did not have a longer-turnaround phase, Site A observed 0 HOCI cases and Sites D and E returned SRT reports for 0/1 and 0/2 HOCI cases, respectively, in this phase.

We did not detect a statistically significant change in weekly incidence of HAIs in the longer-turnaround (incidence rate ratio 1.60, 95%CI 0.85-3.01; *P=*0.14) or rapid (0.85, 0.48-1.50; 0.54) intervention phases in comparison to baseline phase across the 14 sites (Table 3), and incidence rate ratios were comparable in our per-protocol analysis. Similarly, there was only weak evidence for an effect of phase on incidence of outbreaks in both intention-to-treat and per-protocol analyses, with wide confidence intervals inclusive of no difference in incidence (Table 3).

**Table 3:** Incidence outcomes by study intervention phase, overall and within the 7/14 sites included in the ‘per protocol’ sensitivity analysis

|  | Study phase |  |  | IRR† (95% CI, P) |  |
| --- | --- | --- | --- | --- | --- |
|  | Baseline | Longer-turnaround | Rapid | Longer-turnaround<br>vs<br>baseline | Rapid vs<br>baseline |
| *All sites* |  |  |  |  |  |
| *n* HOCl cases | 850 | 373 | 947 | — | — |
| *n* IPC-defined HAIs | 488 | 207 | 576 | — | — |
| Weekly inc. of IPC-defined HAIs per 100 inpatients, mean (median, IQR, range)* [primary outcome] | 1.0 (0.5, 0.0-1.4, 0.0-5.6) | 0.7 (0.3, 0.0-0.7, 0.0-7.6)‡ | 0.6 (0.3, 0.0-0.8, 0.0-5.3)‡ | 1.60 (0.85-3.01; 0.14) | 0.85 (0.48-1.50; 0.54) |
| *n* IPC-defined outbreak events | 129 | 33 | 114 | — | — |
| Weekly inc. of IPC-defined outbreak events per 100 inpatients, mean (median, IQR, range)* | 0.3 (0.1, 0.0-0.4, 0.0-2.3) | 0.1 (0.0, 0.0-0.1, 0.0-0.9)‡ | 0.1 (0.0, 0.0-0.0, 0.0-0.9)‡ | 1.09 (0.38-3.16; 0.86) | 0.58 (0.24-1.39; 0.20) |
| *n* IPC+sequencing-defined outbreak events | — | 40 | 133 | — | — |
| Weekly inc. of IPC+seq. -defined outbreak events per 100 inpatients, mean (median, IQR, range)* | — | 0.1 (0.0, 0.0-0.1, 0.0-1.3)‡ | 0.1 (0.0, 0.0-0.1, 0.0-0.8)‡ | — | — |
| *Sensitivity analysis* |  |  |  |  |  |
| *n* HOCl cases | 290 | 143 | 533 | — | — |
| *n* IPC-defined HAIs | 179 | 91 | 337 | — | — |
| Weekly inc. of IPC-defined HAIs per 100 inpatients, mean (median, IQR, range)* [primary outcome] | 0.3 (0.0, 0.0-0.3, 0.0-3.0) | 0.3 (0.0, 0.0-0.0, 0.0-3.4)‡ | 0.4 (0.0, 0.0-0.3, 0.0-5.3)‡ | 2.21 (0.82-5.92; 0.10) | 1.75 (0.75-4.08; 0.16) |
| *n* IPC-defined outbreak events | 58 | 14 | 55 | — | — |
| Weekly inc. of IPC-defined outbreak events per 100 inpatients, mean (median, IQR, range)* | 0.1 (0.0, 0.0-0.1, 0.0-1.3) | 0.0 (0.0, 0.0-0.0, 0.0-0.6)‡ | 0.0 (0.0, 0.0-0.0, 0.0-0.9)‡ | 0.83 (0.14-4.93; 0.80) | 0.46 (0.11-1.86; 0.21) |
| *n* IPC+seq.-defined outbreak events | — | 14 | 67 | — | — |
| Weekly inc. of IPC+seq. -defined outbreak events per 100 inpatients, mean (median, IQR, range)* | — | 0.0 (0.0, 0.0-0.0, 0.0-0.6)‡ | 0.1 (0.0, 0.0-0.0, 0.0-0.7)‡ | — | — |

HAI, hospital-acquired infection; HOCl, hospital onset COVID-19 infection; IPC, infection prevention and control; IQR, interquartile range; seq., sequencing.

We compared HOCI-level impacts of the sequence report between phases. Nosocomial linkage not suspected by IPC was identified in 6.7% and 6.8% of all HOCI cases in the rapid and longer-turnaround phases, respectively (OR for ‘rapid vs longer-turnaround’ 0.98, 95%CI 0.46-2.08; *P*=0.95) (Table 2) and in 11.4% and 12.6% respectively of cases where the report was returned. For 25 cases in the rapid and five cases in the longer-turnaround phase phylogenetic trees were used for sequences with <90% genome coverage, with three from the rapid phase showing previously unidentified linkage.

IPC practices were changed in 7.8% and 7.4% of all HOCI cases in the rapid and longer-turnaround phases, respectively (OR for ‘rapid vs longer-turnaround’ 1.07, 0.34-3.38; *P*=0.90) and 17.2% and 11.6% respectively of cases where the report was returned. No one specific change to IPC action dominated those recorded (Table S2). When restricted to higher performing sites (i.e. per-protocol), IPC practice was changed in a greater proportion of all HOCI cases in the rapid (9.9%) in comparison to the longer-turnaround (0.7%) sequencing phase (OR for ‘rapid vs longer-turnaround’ 15.55, 1.30-1.85; *P*=0.01) and 16.7% and 1.1% respectively of cases where SRT reports were returned. The impact of phase on detecting nosocomial linkage was similar.

IPC teams more commonly reported finding the sequence reports useful in the rapid sequencing, 303/428 (70.8%) compared to the longer-turnaround phase, 107/215 (49.8%) (OR 0.82 rapid *vs* longer-turnaround, 0.12-5.46; *P*=0.82), and the difference was more pronounced in the per-protocol analysis (79.0 vs 27.2%, respectively; OR 3.44, 0.28-42.61; *P*=0.41). We explored this association further using the actual time to return of the reports (Figure 2). In the per-protocol analysis an impact on IPC actions was observed in 20.7% (45/217) of HOCI cases in which the SRT report was returned within 5 days, but in very few cases beyond this, with this trend less apparent when data from all sites were considered. Figure 2 also displays a strong decline in reported usefulness of the SRT with increasing turnaround time, both across all sites and in the per-protocol analysis. Sequence reports were considered useful in 79.1% (182/230) of cases if returned within 5 days for all sites (169/216, 78%, in per protocol analysis).

**Figure 2.**
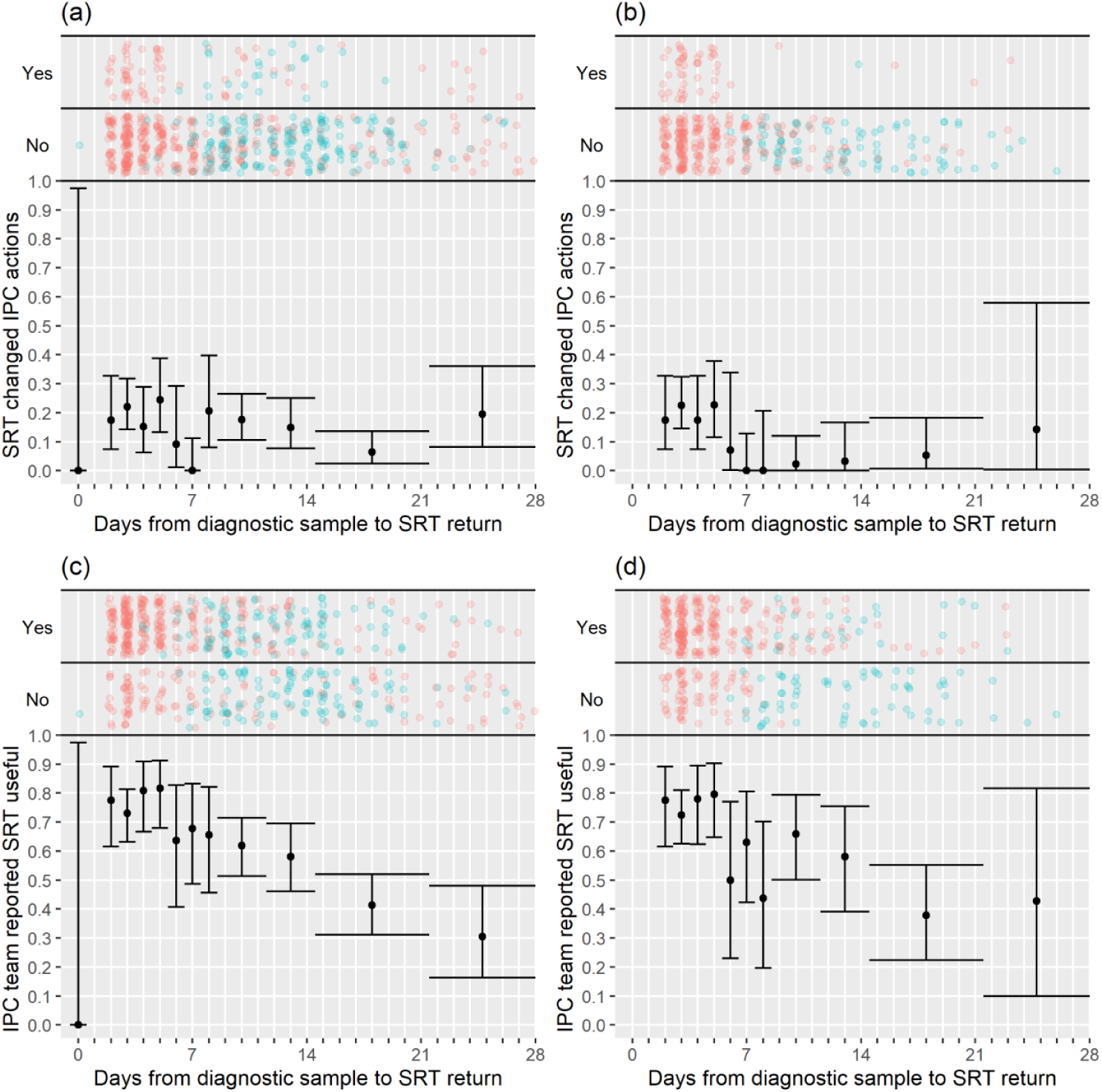
Plots of the proportion of returned SRT reports that had an impact on IPC actions ((a) and (b)) and that were reported to be useful by IPC teams ((c) and (d)). Data are shown for all sites in (a) and (c), and for the seven sites included in the ‘per protocol’ sensitivity analysis in (b) and (d). Results are only shown up to turnaround times of ≤28 days, and grouped proportions are shown for ≥9 days because of data sparsity at higher turnaround times. Error bars show binomial 95% CIs. “Yes” and “No” outcomes for individual HOCI cases are displayed, colour-coded by rapid (red) and longer-turnaround (blue) intervention phases and with random jitter to avoid overplotting. “Unsure” responses were coded as “No” for (c) and (d).

SRT reports suggested that 91.3% of HOCI patients had acquired their infection post-admission (580/635, Table 2). In 91.9%, (589/641, Table S2) of cases the reports were interpreted as supportive of IPC actions already taken. SRT reports also suggested post-admission infection in the majority of indeterminate HAIs (diagnosed 3-7 days from admission) (176/223, 78.9%).

Our analysis models reveal important findings beyond the effect of the intervention. The analysis model for the incidence of HAIs identified independent positive associations with the proportion of current SARS-CoV-2 positive inpatients, the local community incidence of new SARS-CoV-2 cases (which peaked in December 2020 to January 2021, Figure S2 and S3), and calendar time (modelled as ‘study week’). Adding the proportion of local community cases that were Alpha (lineage B.1.1.7) variant did not lead to a statistically significant improvement in model fit (*P*=0.78). The observed weekly HOCI incidence rates varied substantially from 0 to 7.6 per 100 SARS-CoV-2 negative inpatients, with peaks aligning with those for local community incidence (Figure S2).

From modelling outbreaks, positive associations were similarly found for both hospital prevalence and community incidence of SARS-CoV-2 (Figure S4). The median number of HOCIs per IPC-defined outbreak event was four, with the largest observed outbreak including 43 HOCIs (Table S1).

Extensive qualitative analyses^[15]^ found high levels of acceptability for the SRT sequencing reports, which supported decision-making about IPC activity (e.g. stand down some IPC actions or continue as planned). In several sites the major barriers to embedding and normalising the SRT within existing systems and processes were overcome. The SRT did provide new and valued insights into transmission events, outbreaks and wider hospital functioning but mainly acted to offer confirmation and reassurance to IPC teams. Critically, the capacity to generate and respond to these insights effectively on a case-by-case basis was breached in most sites by the volume of HOCIs, and the limits of finite human and physical resource (e.g. hospital layout).

### Health economics findings

The analysis of the SARS-CoV-2 genome sequencing in the 10 laboratories who performed the tests for the sites included in the study showed that mean per sample costs were on average higher for rapid (£78.11) versus longer-turnaround (£66.94) sequencing. (Table S4). Consumables were the highest cost driver of the sequencing process accounting for 66% in rapid and 67% in longer turnaround sequencing.

Several factors affected the costs of genome sequencing. There was a general tendency of increasing returns to scale, with average per-sample costs of genome sequencing tending to decrease as the batch size increases; cost per sample in reagents also depends highly on how many samples are processed per batch. Another factor was the sequencing platform and protocols used: some processes had been automated which reduced the hands-on input.

## Discussion

This study constitutes the largest prospective multicentre evaluation study of viral whole genome sequencing (WGS) for acute IPC investigation of nosocomial transmission conducted to date. The study was run as part of routine practice within the NHS, and the challenges faced in implementing the intervention reflect the context and barriers at present in the UK. Among sites with the most effective implementation of the sequencing intervention we showed that feedback within 5 days of diagnosis allowed for maximal impact on IPC actions. While the study was not able to show a direct impact of sequencing on the incidence of nosocomial transmission, IPC teams, particularly in the per-protocol analysis, were almost all positive in their perception of the utility of viral sequencing for outbreak investigation.

Outbreak investigations are inherently complex and must take account of uncertainty regarding transmission links, even in the presence of high-quality genomic data^[16]^. Interventions centred on IPC practices often need to be evaluated at the hospital level in order to allow for impacts on transmission across an institution as a whole^[17]^, meaning that large multicentre studies are required to generate high-quality evidence. Standardisation of data collection with complex structures across multiple hospital sites is a considerable challenge. A review of IPC practice guidelines conducted prior to the SARS-CoV-2 pandemic found that most recommendations were based on evidence from descriptive studies, expert opinion and other low-quality evidence^[18]^.

The use of viral WGS for public health surveillance has become firmly established in the UK for SARS-CoV-2^[19]^. This enabled early detection of the increased transmissibility and health impact of the Alpha variant^[20]^ and subsequent monitoring of the Delta^[21]^ and Omicron variants^[22]^. However, whilst viral WGS for acute outbreak investigation has been shown for both SARS-CoV-2^[2, 4, 23, 24]^ and other viruses^[25-27]^ to better identify sources of hospital acquired infections and transmission chains, its impact on the management and outcome of nosocomial infection has not previously been quantified. Our study provides a substantial body of evidence regarding the introduction of viral WGS into hospital functioning, routine IPC practice, its potential impact on outbreak management and the challenges that need to be overcome to achieve implementation across the UK.

There are several limitations that may have impacted on the results from this study. The study was conducted between October 2020 and April 2021. In this period, the local community incidence for the study sites ranged from <50 to >1200 weekly cases per 100,000 people. There were corresponding large variations in the healthcare burden of COVID-19, with several sites recording weeks when more than half of all inpatients were SARS-CoV-2 positive. High community infection rates and associated increases in the incidence of HOCI cases contributed to difficulties for site research teams in generating good quality viral sequences and reports for all HOCI cases within target timeframes. It may therefore be more achievable to develop systems for rapid viral WGS and feedback for endemic respiratory viruses at lower and more consistent levels. The peak in SARS-CoV-2 levels in December 2020 to January 2021 corresponded to the rise of the highly transmissible Alpha variant in the UK^[20]^. We did not find that the local prevalence of the Alpha variant was associated with the incidence rate of HAIs, beyond any effect mediated by higher community incidence. This matches the conclusions of a previously reported sub-study analysis using data from our sites^[28]^.

The study intervention made use of a bespoke sequence reporting tool^[10]^. The SRT combined both patient-meta-data and sequencing data, providing a single-page, easily interpretable report for IPC teams. It also facilitated standardisation of data collection across sites. Interestingly, while HOCIs diagnosed 3-7 days after admission are generally excluded from assessments of nosocomial SARS-CoV-2 infections^[8]^, because of difficulty in distinguishing them from community-acquired infections, the SRT reported the majority (78.9%) of these indeterminate HAIs as being hospital-acquired. This confirms findings from a retrospective study using genomic linkage^[23]^, and may reflect a shorter incubation time for the Alpha variant compared to earlier variants^[29]^, indicating that definitions used for monitoring and reporting may need to be kept under active review.

A number of limitations of the SRT were recognised, and work is ongoing to rectify these for future studies. The SRT’s probability calculations did not include patient and healthcare worker (HCW) movements. The SRT gave feedback on cases that could plausibly form part of the same outbreak but did not identify direct transmission pairs or networks, as has been done in other studies^[16, 30]^. Finally, samples with less than 90% genome coverage were not included within the reporting system, despite the fact that they may still be useful for phylogenetic analyses.

The study sites varied in their ability to process sequence and meta-data and generate and distribute reports in a timely manner (Figure 1). Sites that had established teams with existing genomics expertise and on-site sequencing facilities were generally more successful at implementing the SRT into clinical practice^[15]^. There is a need to focus on how sequencing and reporting processes can be integrated within local infrastructure and tailoring of local processes to ensure clear chains of communication from diagnostic labs through to the IPC team. Precisely understanding the barriers to achieving rapid turnaround times is key to future IPC use of viral WGS and is currently being analysed in a follow-up secondary analysis. Standardising and automating more of the SRT production pipeline will also help reduce the implementation burden at sites.

The study covered a period in which a national vaccination programme was initiated for HCWs and the elderly population in the UK, commencing with those ≥80 years from 8 December 2020. We had planned to include data on the proportion of HCWs who had received at least one vaccine dose as a variable in the analysis of incidence outcomes. This was subsequently not included because data was only available from 10 sites, for which rollout of HCW vaccination was broadly consistent. As such, any effect of HCW or patient vaccination on the incidence outcomes would form part of the estimated association with calendar time.

With the sequencing technology now available and high levels of interest in viral genomics for public health, there is the potential to incorporate viral WGS into routine IPC practice. Many publications have already highlighted the utility of viral sequence data for changing IPC policy and auditing the management of outbreaks^[2, 4, 23-27]^. Our study provides the first evidence that with faster turnaround times, viral sequences can inform ongoing IPC actions in managing nosocomial infections; results returned within ≤5 days from sampling to result changed the actions of IPC teams in around 20% of cases. The SRT, by rapidly combining sequence and patient meta-data, was also better able than standard IPC definitions alone to distinguish hospital and community acquired infections within a clinically relevant time scale. The difference in the cost of rapid compared with longer-turnaround hospital sample sequencing is low relative to the overall cost level at present (Table S4). Assuming SARS-CoV-2 sequencing for public health purposes continues, the added cost of rapid sequencing for IPC purposes could potentially be offset by the benefits accrued.

While we did not show an impact of sequencing on the numbers of HAIs or outbreaks, the evidence that these correlated with the high community SARS-CoV-2 rates suggests that factors beyond the control of IPC were influential. Our study nonetheless provides valuable evidence regarding the implementation and utility of this technology for IPC, and potentially will have a greater positive impact on IPC practice outside of the burdens imposed by a pandemic. Importantly for future research, we provide a wealth of data on why the study worked better at some sites than others, and the challenges that would need to be overcome to make full use of viral genome sequencing for IPC practice more widely.

## Supporting information

HOCI Investigator list

COG-UK Consortium list

CONSORT checklist

## Data Availability

Fully anonymised versions of the datasets generated and analysed during the current study will be stored on a data repository and sharing platform CSDR (https://clinicalstudydatarequest.com/) so that the data may be reused by other researchers. This will include individual participant level data, data dictionaries and the statistical analysis plan for this study. This will be done within 6 months of public reporting of results. Access through the data sharing platform requires submission of a viable research plan for review.

## Acknowledgments

COG-UK is supported by funding from the Medical Research Council (MRC) part of UK Research & Innovation (UKRI), the National Institute of Health Research (NIHR) [grant code: MC_PC_19027], and Genome Research Limited, operating as the Wellcome Sanger Institute.

We also acknowledge the support of the independent members of the Joint Trial Steering Committee and Data Monitoring Committee (TSC-DMC): Prof Marion Koopmans (Erasmus MC), Prof Walter Zingg (University of Geneva), Prof Colm Bergin (Trinity College Dublin), Prof Karla Hemming (University of Birmingham), Prof Katherine Fielding (LSHTM). As well as TSC-DMC non-independent members: Prof Nick Lemoine (NIHR CRN), Prof Sharon Peacock (COG-UK). We would also thank members of COG-UK who have directly supported the study: Dr Ewan Harrison (Cambridge University), Dr Katerina Galai (PHE), Dr Francesc Coll (LSHTM), Dr Michael Chapman (HDR-UK), Prof Thomas Connor and team (Cardiff University), Prof Nick Loman and team (University of Birmingham). We also thank the COG-UK Consortium, and the UK National Institute for Health Research Clinical Research Network (NIHR CRN).

## Declaration of interests

The authors do not have any declarations of interest.

## Contributors

JBr, JBl, ACo, OS, MP, GN, DGP, MP, JP, CP, LBS, TIdS, PF and FM planned the study design and wrote the Protocol. AH, NM, GYS, TM, KS, TS, YT, TM, CP, ACh, MTCM, ET, GN and TIdS were clinical principal investigators at each site. RW, AD, DR, EN, SR, DS, KL, IM, BK and SH were heads of clinical and academic sequencing teams. JH, FF, SH, RG, MB, ML, MP, AW, SR, AT, FC and MC conducted bioinformatics and data processing at each site or in relation to the study as a whole. SP is Executive Director of the COG-UK consortium, coordinating the UK national sequencing program. JBl and AMN coordinated site activities and data management. OS and ACo wrote the statistical analysis plan. OS coded the primary analysis, checked by ACo. OTS and ACo had full access to and verified the final analysis dataset. OS, JBr, JBl, FM, PF, MP and ACo drafted the initial version of this manuscript. All authors reviewed the final manuscript and approved this for submission.

## Supplementary Appendix

### Methods

#### Sample size estimation

There was uncertainty in the number of HOCIs that would be identified at each site during each of the intervention periods, with the rapid sequencing phase being 8 weeks’ duration. We assumed there may be an average of 10 HOCIs/week per site during this intervention period, a total of 80 per site. Within a typical site this would allow us to estimate the proportion of HOCIs with genotypic linkage to another case(s) not detected by IPC processes with minimum precision of +/-9.4%. Similarly we would be able to estimate the proportion of HOCIs where an action is taken that would not have occurred without sequencing within +/-9.4%, with a pooled estimate of key proportions across the 14 sites implementing rapid sequencing within +/-6.5% assuming an intra-cluster correlation coefficient of 0.05.

Comparing the proportion of HOCIs with genotypic linkage to another case(s) not detected by IPC processes between rapid testing and delayed testing phases across all sites, the study aimed for at least 80% power to detect a percentage point difference of 11% (two-sided test with alpha=0.05, considering proportions of 55.5% vs 44.5% which would be associated with minimum power for a difference of this magnitude).

For the outcome of weekly incidence of IPC-defined HOCIs, using an approximate Normal distribution for weekly counts there was 86.7% power to demonstrate a reduction from 12 IPC-defined HOCIs per week in the baseline phase to 10 per week during the rapid testing phase across all sites, under 5% significance level two-tailed testing. However, these calculations correspond to a variance of 12 for weekly counts based on the Poisson distribution, but the presence of over-dispersion of weekly counts would lead to a lower power to detect a difference. Using an overdispersion parameter of 0.82 based on retrospective analysis of data from Sheffield and Glasgow (dataset as described by Stirrup et al.^[10]^) resulted in 81% power to detect a reduction in mean weekly incidence from 12.5 to 10.

##### Planned secondary outcomes dropped from formal analysis

We did not carry out formal statistical analysis for the following planned secondary outcomes:

- Weekly incidence of IPC+sequencing-defined SARS-CoV-2 hospital outbreaks, measured as incidence rate per week per 100 non-COVID-19 inpatients, during each phase of the study based on case report forms.
- The number of healthcare worker (HCW) periods of sickness/self-isolation as assessed as a proportion of the number of staff usually on those wards impacted by HOCI cases, for all phases of the study.

The first of these was dropped because of incomplete sequencing coverage of HOCI cases in the intervention phases – it was not felt that this would add useful information given the level of sequencing achieved and the null results for other incidence outcomes. The second was dropped because of low levels of data completion at most of the study sites.

We also omitted specific reporting of the secondary outcome of ‘Ideal changes to IPC actions following receipt of sequencing report’. This was because no recommended changes to IPC actions were recorded that were also recorded as ‘not implemented’. As such, this outcome was identical to ‘changes to IPC actions following receipt of sequencing report’.

#### Coding of primary and secondary outcomes Primary outcome 1

##### Incidence of IPC-defined SARS-CoV-2 HAIs

In order to standardise this measure across sites, ‘IPC-defined SARS-CoV-2 HAIs’ were considered to be all HOCIs with an interval of ≥8 days from admission to symptom onset (if known) or sample date (i.e. those meeting the PHE definition of a probable or definite HAI^[13]^). Incidence was expressed ‘per 100 non-COVID-19 inpatients per site per week’, and was evaluated for study baseline and intervention phases.

#### Primary outcome 2

##### Identification of SARS-CoV-2 nosocomial transmission using sequencing data

For each HOCI case during the intervention phases, the occurrence of this outcome was defined as positive where the following two answers had been recorded in the Hospital Transmission section of the relevant clinical reporting form (CRF04):

> “Is sequencing report suggestive that patient is part of a hospital outbreak (i.e. involving ≥2 patients or HCWs in the hospital)?: Yes”
>
> &
>
> “If yes, was linkage to one or more of these patients suspected at initial IPC investigation?:
>
> No”

The occurrence of this outcome was considered to be negative if the following answer was recorded:

> “Is sequencing report suggestive that patient is part of a hospital outbreak (i.e. involving ≥2 patients or HCWs in the hospital)?: No”

Or if the following combination was recorded:

> “Is sequencing report suggestive that patient is part of a hospital outbreak (i.e. involving ≥2 patients or HCWs in the hospital)?: Yes”
>
> &
>
> “If yes, was linkage to one or more of these patients suspected at initial IPC investigation?: Yes”

The outcome will be considered missing if either the first question was not answered, or if the first question was answered ‘Yes’ and the second question was not answered or was answered ‘unknown’.

The outcome was also be considered negative if the viral sequence and sequence report had not been returned during the period of study data collection.

This outcome was only be evaluated for study sequencing intervention periods.

#### Secondary outcome 1

##### Incidence of IPC-defined SARS-CoV-2 hospital outbreaks

An IPC-defined SARS-CoV-2 hospital outbreak was defined as the occurrence of at least two HOCI cases on the same ward, with at least one having an interval of ≥8 days from admission to symptom onset (if known) or sample date, and with the outbreak event considered to be concluded if there was a gap of 28 days before the observation of another HOCI case^[13]^. This was evaluated using the ward location recorded at patient registration into the study (CRF01) for HOCI cases, cross-checked against patient movement data to confirm location at diagnostic sampling. Outbreak events were considered to have occurred on the date of diagnosis of the first HOCI case. This outcome was be evaluated for study baseline and intervention phases.

#### Secondary outcome 2

##### Changes to IPC actions following receipt of sequencing report

For each HOCI case, the occurrence of this outcome was defined as positive if ‘Yes’ is the answer to either of the following two questions in the ‘Sequencing report impact on IPC team’ section of CRF04:

> “Overall, did the sequencing report change IPC practice for this ward?”
>
> And/or
>
> “Has the sequencing report information been used in IPC decisions beyond this patient’s ward?”

And/or, if any specific changes to IPC practice were recorded on CRF04.

The occurrence of the outcome was considered negative if at least one of these questions was answered ‘No’ and neither is answered ‘Yes’, and it was considered missing if neither were answered.

This outcome was only evaluated for study sequencing intervention periods.

#### Secondary outcome 3

##### Ideal changes to IPC actions following receipt of sequencing report

A binary outcome was defined for each HOCI patient. This was based on the value of Secondary Outcome 3, but was additionally be defined as positive (whether Secondary Outcome 3 was negative or missing) if an ‘increase’ or ‘decrease’ that was not implemented was recorded for any of the actions in the ‘other recommended changes to IPC protocols’ section of CRF04.

This outcome was only evaluated for study sequencing intervention periods.

#### Secondary outcome 4

##### Incidence of IPC+sequencing-defined SARS-CoV-2 hospital outbreaks

An IPC+sequencing-defined SARS-CoV-2 hospital outbreak was defined as the occurrance of at least two HOCI cases on the same ward that form a genetic cluster with maximum viral sequence pairwise SNP distance of 2 between each individual included and their nearest neighbour within the cluster. This was evaluated using the ward location recorded at patient registration into the study (CRF01), with HOCI cases sorted into outbreak groups using the lists of close sequence matches on unit-ward as returned by the SRT and recorded in CRF03.

Outbreak events were considered to have occurred on the date of diagnosis of the first HOCI case. This outcome was evaluated for study sequencing intervention periods for all sites.

#### Secondary outcome 5

##### HCW sickness

The proportion of HCWs on sick leave due to COVID-19 was calculated using the ‘Current staffing levels on ward’ section of CRF02. Analysis was performed using the first available data within each IPC-defined SARS-CoV-2 hospital outbreak (as per secondary outcome 1), so as to provide a measure of the level of staff absence at the start of each outbreak. This outcome was evaluated for study baseline and intervention phases.

#### Changes with respect to the statistical analysis plan (SAP)

It was planned that the cumulative proportion of HCWs vaccinated at each site for each study week would also be included as a covariate for the analysis models of incidence outcomes. However, this was dropped because these supplementary data could not be obtained from four sites (and one site was only able to provide partial data). Where available, the data showed the rollout of HCW vaccination to be broadly consistent across sites. As such, any effect of HCW vaccination on the incidence outcomes would be incorporated into estimates of variation in relation to calendar time.

#### Small sample correction

The topic of small sample corrections for cluster randomised and other cluster-structured studies (e.g. stepped wedge trials) with outcomes that are not normally distributed is an area of ongoing active research. To our knowledge, there do not exist any studies regarding appropriate corrections for clustered data when analysing an outcome with negative binomial distribution. However, when calculating P-values and confidence intervals for the primary and secondary outcomes we will use a *t*-distribution with 12 or 13 degrees of freedom (*n* clusters – *n* relevant parameters) in order to ensure that there is not an inflated type-1 error rate. This correction has shown appropriate characteristics in simulation studies of analyses of binary outcomes using mixed effects models and generalised estimating equations^[31, 32]^.

#### Decision regarding continuation of study into final phase

A decision regarding the final phase of the study (Period 4) was planned for April 2021, with the options being: ending of the study at Period 3, a further phase of rapid sequencing at each site or a further phase of ‘baseline’ date collection without use of the SRT. A recommendation regarding this decision was made by the study investigators and agreed with the TSC-DMC. The decision was determined by the course of the epidemic and the progress of vaccination among key risk groups, and by the quantity of data collected by the end of Period 3. The decision was not based on any interim evaluation of the effect of the sequencing intervention under investigation on the incidence of nosocomial infection.

A decision was made to stop the study at the end of Period 3 because the total sample size was close to that projected for the study, and few new HOCI cases were being recorded at this point in time.

### Results

**Figure S1:**
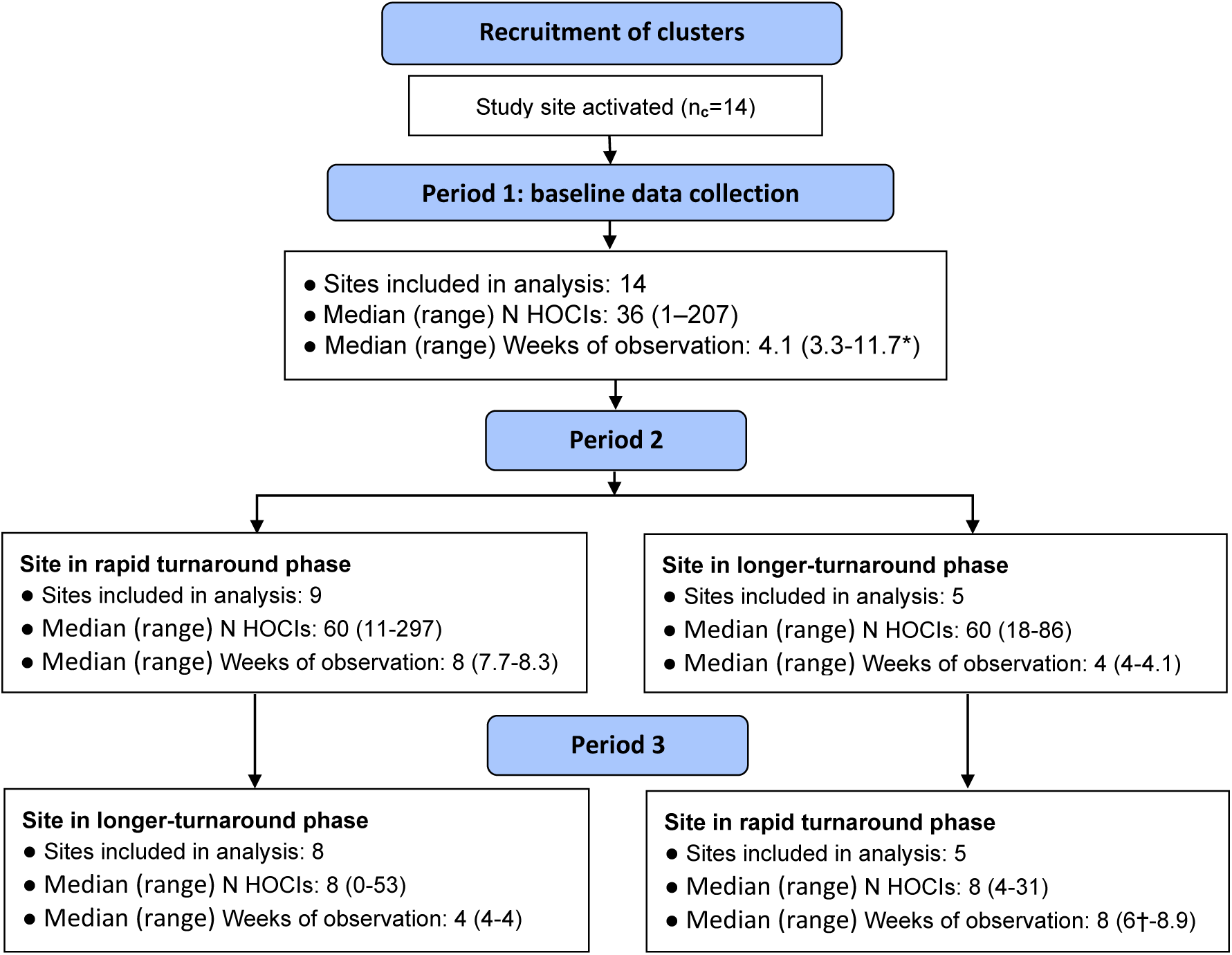
Flow diagram of study site enrolment and intervention implementation *Baseline phase extended for one site due to a complete lack of HOCI cases during first few weeks of study period and omission of longer-turnaround sequencing phase. †Rapid sequencing phase truncated at one site due to cessation of enrolment at all sites.

**Figure S2:**
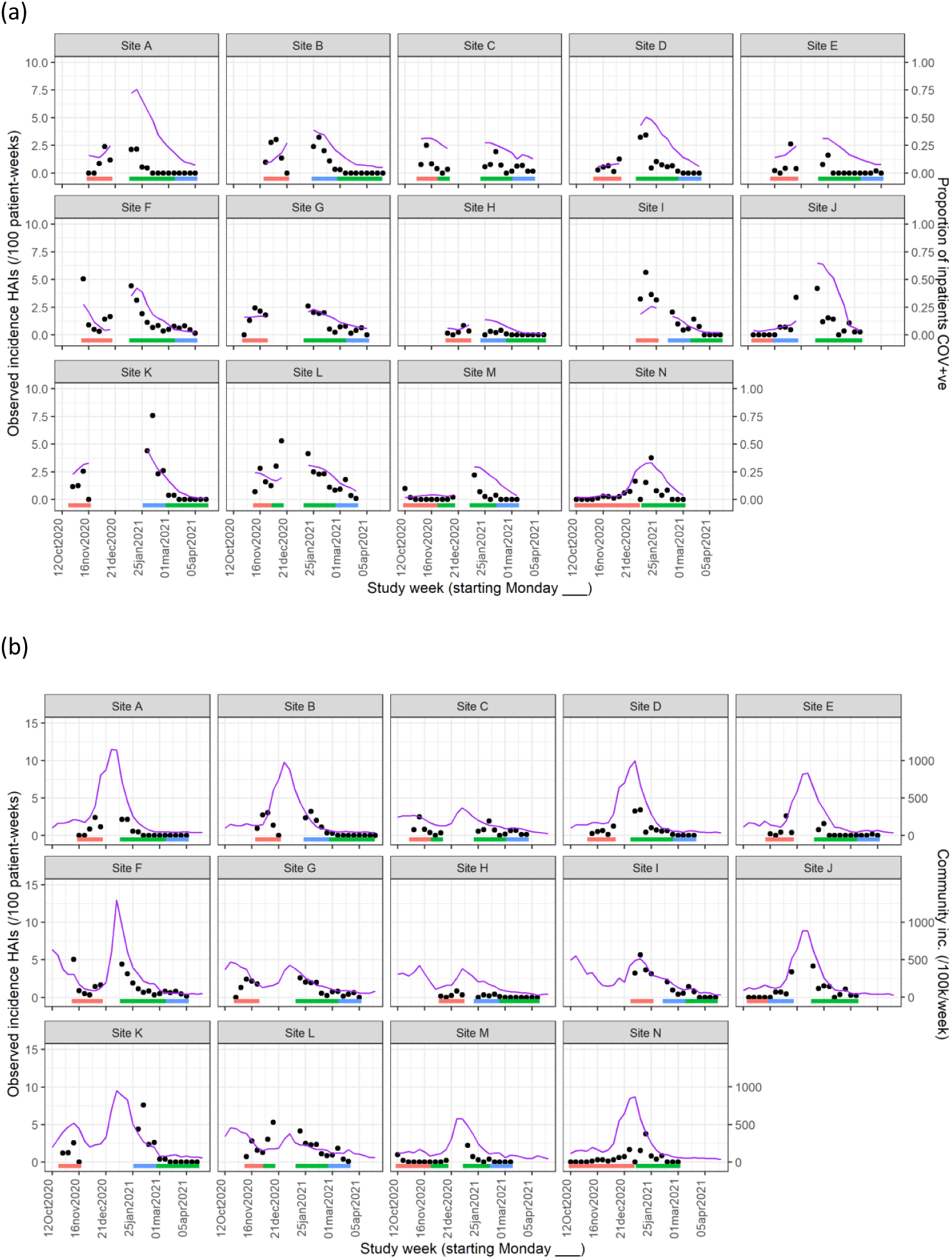
Weekly incidence of HAIs at each site (●), with (a) proportion of all inpatients SARS-CoV-2 +ve and (b) local community incidence of SARS-CoV-2 +ve tests also plotted on the y-axis (purple line). Horizontal bars show the duration of study phases (red: baseline; blue: longer turnaround; green: rapid).

**Figure S3:**
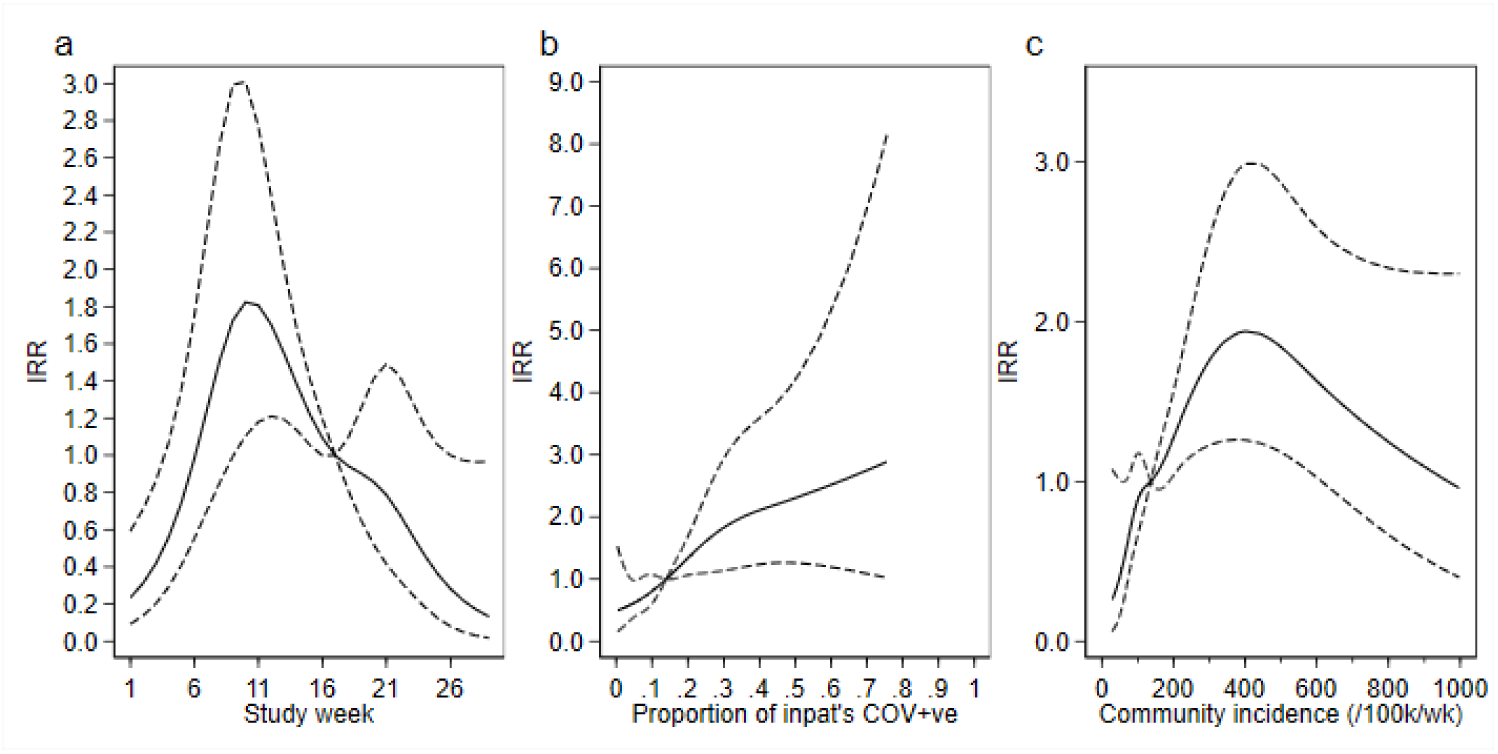
Adustment variables for analysis of weekly incidence of IPC-defined HAIs per 100 inpatients, as described in Table 3. Incidence rate ratios are displayed relative to the median for (a) calendar time expressed as study week from 12^th^ October 2020, (b) proportion of inpatients with positive SARS-CoV-2 test and (c) local community incidence of SARS-CoV-2 (government surveillance data weighted by total set of postcodes for patients at each site). Adjustment for (c) was not pre-specified in the SAP, but adding this variable to the model was associated with a statistically significant improvement in fit (*P* =0.01). The proportion of community-sampled cases in the region that were found to be the Alpha variant on sequencing was also considered, but adding this as a linear predictor did not lead to a statistically significant improvement in model fit (*P*=0.78).

**Figure S4:**
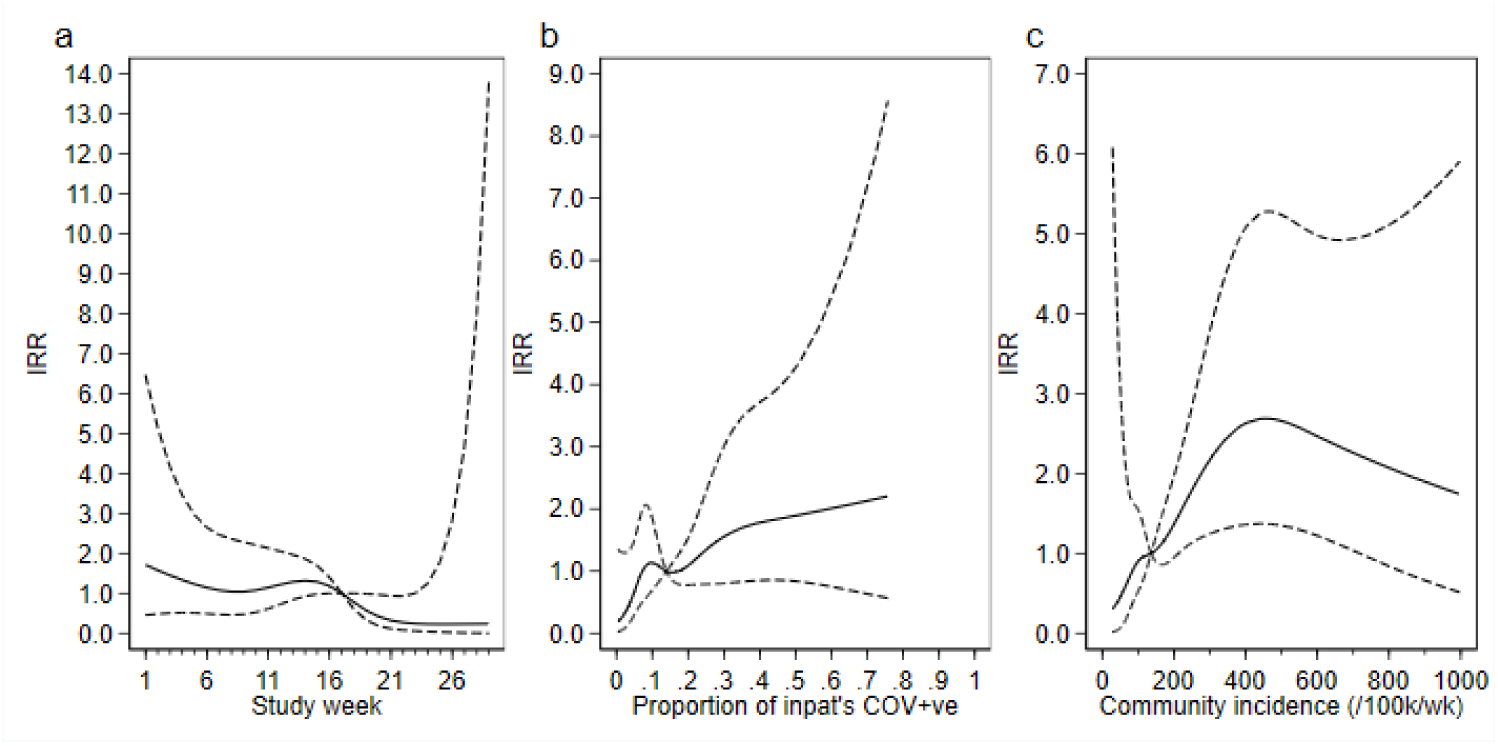
Adustment variables for analysis of weekly incidence of IPC-defined outbreak events per 100 inpatients, as described in Table 3. Incidence rate ratios are displayed relative to the median for (a) calendar time expressed as study week from 12^th^ October 2020, (b) proportion of inpatients with positive SARS-CoV-2 test and (c) local community incidence of SARS-CoV-2 (government surveillance data weighted by total set of postcodes for patients at each site). Adjustment for (c) was not pre-specified in the SAP, but adding this variable to the model was associated with a near-statistically significant improvement in fit (*P*=0.05) and was included for consistency with the analysis of individual HAIs. The proportion of community-sampled cases in the region that were found to be the Alpha variant on sequencing was also considered, but adding this as a linear predictor did not lead to a statistically significant improvement in model fit (*P*=0.80).

**Figure S5:**
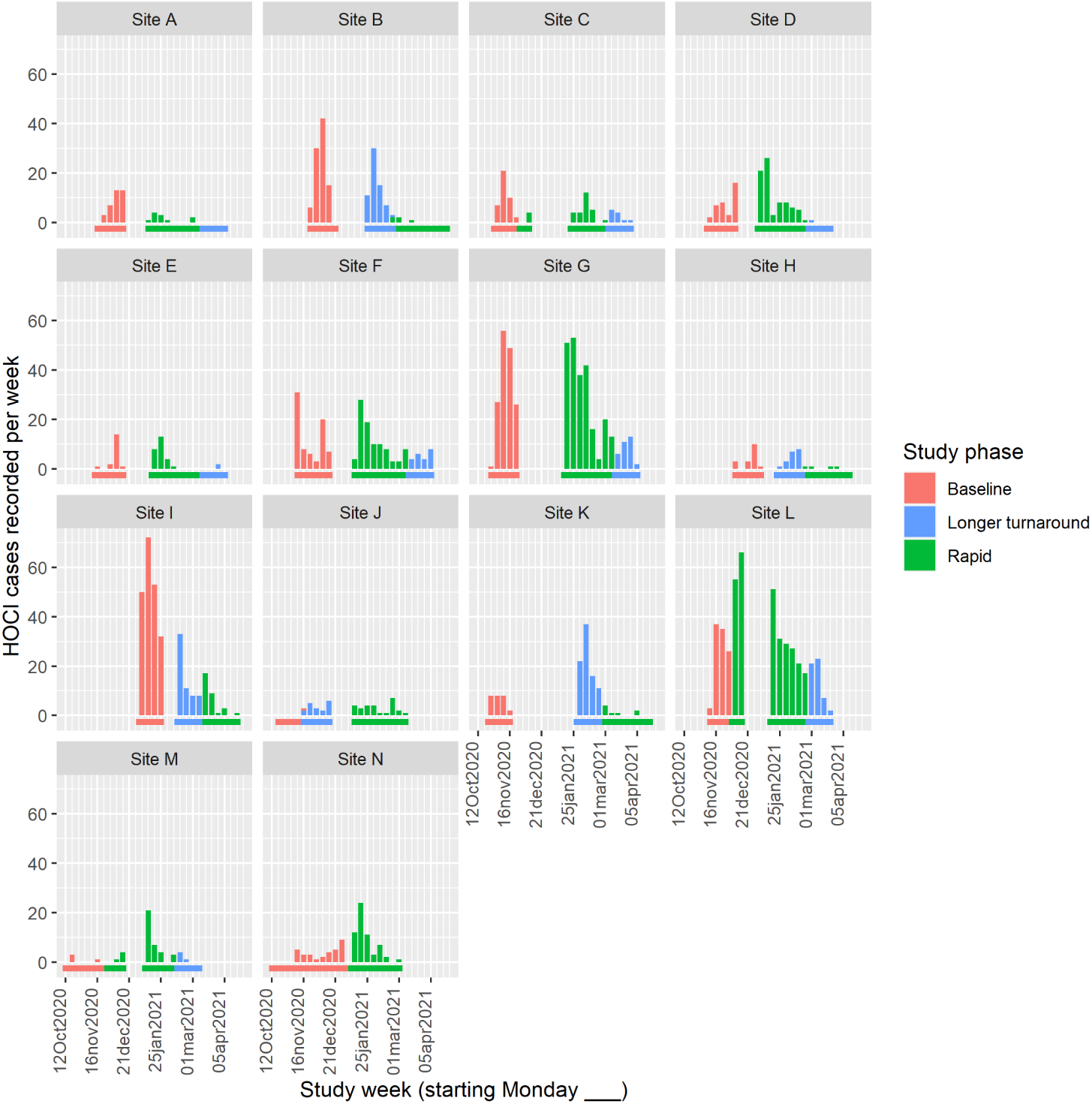
Weekly counts of enrolled HOCI cases by date of positive test return for each site, color-coded by intervention phases. Horizontal bars show the duration of study phases.

**Table S1:** Per outbreak event outcomes by study intervention phase HCW, healthcare worker.

|  | Study phase |  |  |  |
| --- | --- | --- | --- | --- |
|  | Baseline | Longer-turnaround | Rapid | Total |
| **IPC-defined outbreak events** |  |  |  |  |
| *n* outbreak events | 129 | 33 | 114 | 276 |
| *n*/*N* (%) of HOCI cases part of outbreak event | 682/850 (80.2) | 314/373 (84.2) | 763/947 (80.6) | 1759/2170 (81.1) |
| Number of HOCIs per outbreak event, median (IQR, range) | 5.0 (3-8, 2-43) | 5.0 (3-9, 2-24) | 4.0 (2-7, 2-31) | 4.0 (2-8, 2-43) |
| Prop. HCWs on sick leave due to COVID-19, median (IQR, range) [*n*] | 0.13 (0.00-0.35, 0.00-0.50) [13] | 0.05 (0.00-0.18, 0.00-0.30) [7] | 0.20 (0.08-0.33, 0.00-0.89) [14] | 0.13 (0.00-0.31, 0.00-0.89) [34] |
| **IPC+sequencing-defined outbreak events** |  |  |  |  |
| *n* outbreak events | — | 41 | 135 | 176 |
| *n*/*N* (%) of HOCI cases part of outbreak event | — | 292/373 (78.3) | 705/947 (74.4) | 997/1320 (75.5) |
| Number of HOCIs per outbreak event, median (IQR, range) | — | 5.0 (2-8, 2-23) | 3.0 (2-6, 2-29) | 3.0 (2-7, 2-29) |
| *For first HOCI in outbreak:* |  |  |  |  |
| SRT changed IPC practice, *n*/*N* (%*, 95% CI) | — | 4/41 (10.4, 0-21.0) | 19/133 (14.9, 6.6-23.2) | 23/174 (13.2) |
| SRT changed IPC practice for ward, *n*/*N* (%) | — | 2/35 (5.7) | 6/82 (7.3) | 8/117 (6.8) |
| SRT used in IPC decisions beyond ward, *n*/*N* (%) | — | 2/35 (5.7) | 10/82 (12.2) | 12/117 (10.3) |
| IPC team reported SRT to be useful, *n*/*N* (%) |  |  |  |  |
| Yes | — | 20/35 (57.1) | 51/82 (62.2) | 71/117 (60.7) |
| No | — | 9/35 (25.7) | 15/82 (18.3) | 24/117 (20.5) |
| Unsure | — | 6/35 (17.1) | 16/82 (19.5) | 22/117 (18.8) |
| SRT would ideally have changed IPC practice, *n*/*N* (%*, 95% CI) | — | 4/41 (9.8) | 19/133 (14.3) | 23/174 (13.2) |

HCW, healthcare worker.
\*Estimated marginal value from mixed effects model, not raw %, evaluated on intention-to-treat basis with lack of SRT report classified as 'no'.
Odds ratio of SRT changed IPC practice for 'rapid vs longer-turnaround' phases 1.54 (95% CI 0.37-6.44; *P*=0.52).

**Table S2:**
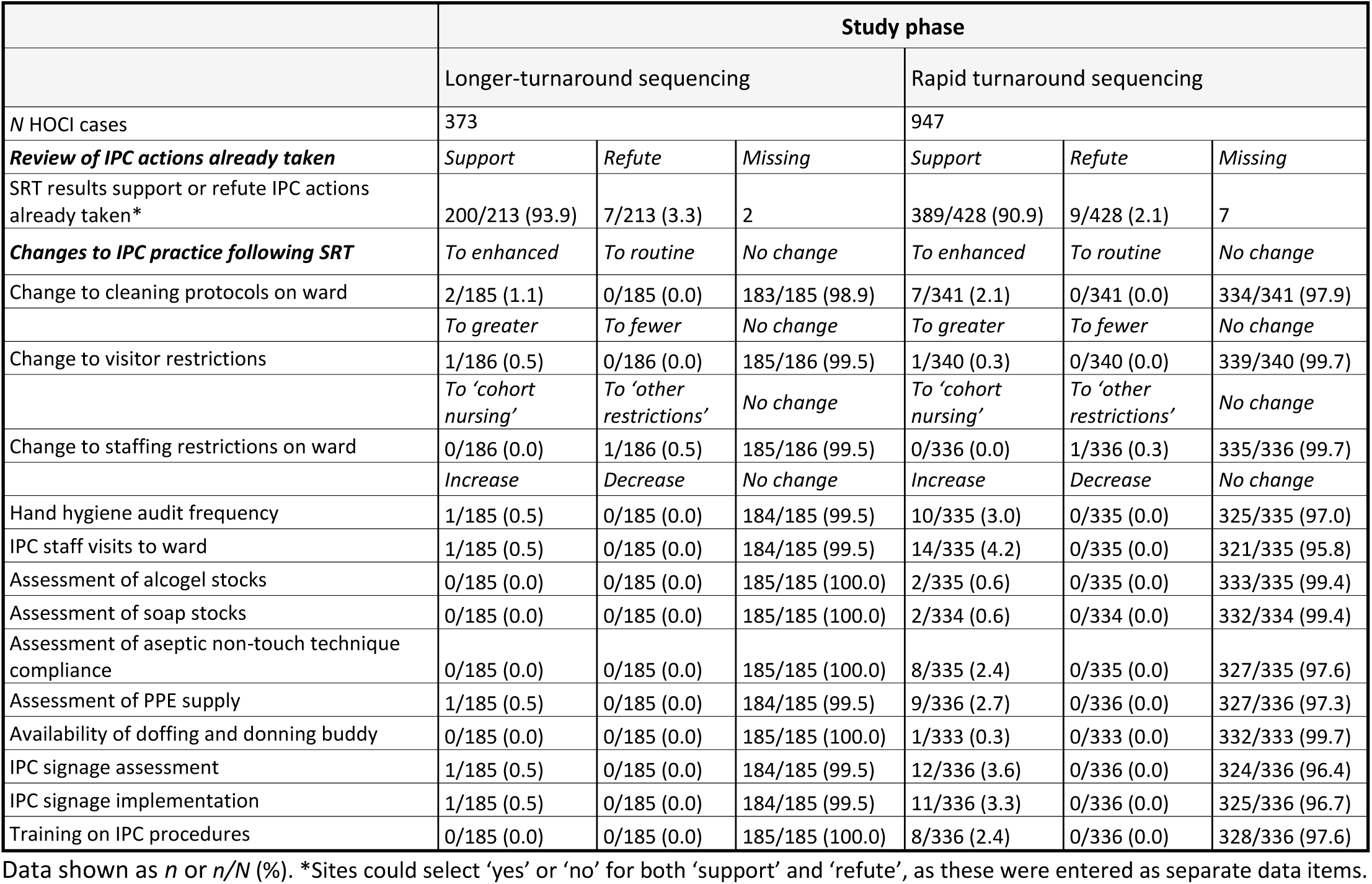
Descriptive summary of impact of sequencing on IPC actions implemented during study intervention phases

**Table S3:** Descriptive summary of impact of sequencing on IPC actions implemented during study intervention phases, only including the first HOCI in each IPC+sequencing-defined outbreak event

|  | Study phase |  |  |  |  |  |
| --- | --- | --- | --- | --- | --- | --- |
|  | Longer-turnaround sequencing |  |  | Rapid turnaround sequencing |  |  |
| *N* HOCl cases | 41 |  |  | 135 |  |  |
| **Review of IPC actions already taken** | *Support* | *Refute* | *Missing* | *Support* | *Refute* | *Missing* |
| SRT results support or refute IPC actions already taken* | 30/35 (85.7) | 3/35 (8.6) | 0 | 71/82 (86.6) | 5/82 (6.1) | 2 |
| **Changes to IPC practice following SRT** | *To enhanced* | *To routine* | *No change* | *To enhanced* | *To routine* | *No change* |
| Change to cleaning protocols on ward | 1/34 (2.9) | 0/34 (0.0) | 33/34 (97.1) | 1/70 (1.4) | 0/70 (0.0) | 69/70 (98.6) |
|  | *To greater* | *To fewer* | *No change* | *To greater* | *To fewer* | *No change* |
| Change to visitor restrictions | 0/34 (0.0) | 0/34 (0.0) | 34/34 (100.0) | 1/70 (1.4) | 0/70 (0.0) | 69/70 (98.6) |
|  | *To 'cohort nursing'* | *To 'other restrictions'* | *No change* | *To 'cohort nursing'* | *To 'other restrictions'* | *No change* |
| Change to staffing restrictions on ward | 0/34 (0.0) | 1/34 (2.9) | 33/34 (97.1) | 0/70 (0.0) | 1/70 (1.4) | 69/70 (98.6) |
|  | *Increase* | *Decrease* | *No change* | *Increase* | *Decrease* | *No change* |
| Hand hygiene audit frequency | 0/34 (0.0) | 0/34 (0.0) | 34/34 (100.0) | 3/70 (4.3) | 0/70 (0.0) | 67/70 (95.7) |
| IPC staff visits to ward | 0/34 (0.0) | 0/34 (0.0) | 34/34 (100.0) | 5/70 (7.1) | 0/70 (0.0) | 65/70 (92.9) |
| Assessment of alcogel stocks | 0/34 (0.0) | 0/34 (0.0) | 34/34 (100.0) | 2/70 (2.9) | 0/70 (0.0) | 68/70 (97.1) |
| Assessment of soap stocks | 0/34 (0.0) | 0/34 (0.0) | 34/34 (100.0) | 2/70 (2.9) | 0/70 (0.0) | 68/70 (97.1) |
| Assessment of aseptic non-touch technique compliance | 0/34 (0.0) | 0/34 (0.0) | 34/34 (100.0) | 3/70 (4.3) | 0/70 (0.0) | 67/70 (95.7) |
| Assessment of PPE supply | 0/34 (0.0) | 0/34 (0.0) | 34/34 (100.0) | 3/70 (4.3) | 0/70 (0.0) | 67/70 (95.7) |
| Availability of doffing and donning buddy | 0/34 (0.0) | 0/34 (0.0) | 34/34 (100.0) | 1/70 (1.4) | 0/70 (0.0) | 69/70 (98.6) |
| IPC signage assessment | 0/34 (0.0) | 0/34 (0.0) | 34/34 (100.0) | 4/70 (5.7) | 0/70 (0.0) | 66/70 (94.3) |
| IPC signage implementation | 0/34 (0.0) | 0/34 (0.0) | 34/34 (100.0) | 4/70 (5.7) | 0/70 (0.0) | 66/70 (94.3) |
| Training on IPC procedures | 0/34 (0.0) | 0/34 (0.0) | 34/34 (100.0) | 4/70 (5.7) | 0/70 (0.0) | 66/70 (94.3) |

Data shown as *n* or *n/N* (%). \*Sites could select 'yes' or 'no' for both 'support' and 'refute', as these were entered as separate data items.

**Table S4:**
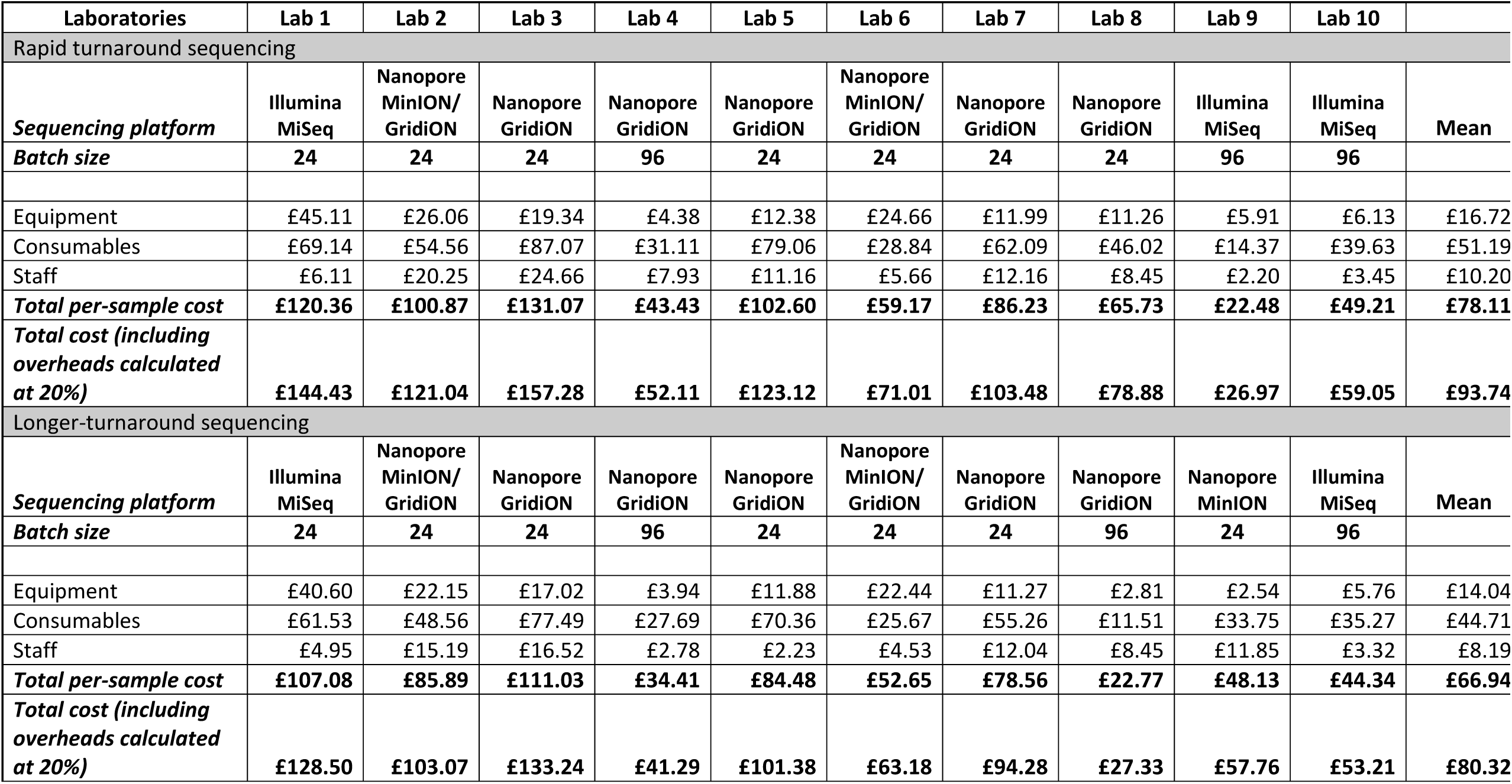
Per-sample costs of SARS-CoV-2 genome rapid and longer turnaround sequencing

## Notes

### Competing Interest Statement

The authors have declared no competing interest.

### Clinical Trial

NCT04405934

### Clinical Protocols

https://www.medrxiv.org/content/10.1101/2021.04.13.21255342v1

### Author Declarations

NHS HRA gave ethical approval for this work (ref: REC 20/EE/0118).

