## Supplementary material for "Evaluating the effectiveness of rapid SARS-CoV-2 genome sequencing in supporting infection control teams: the COG-UK hospital-onset COVID-19 infection study": HOCI Investigator list

### List of COG-UK HOCI Investigators

**Barts site**

| Name of individual | Employing Institution |
|---|---|
| **Teresa Cutino-Moguel** | **Barts Heath NHS Trust** |
| Tabassum Khan | Barts Heath NHS Trust |
| Beatrix Kele | Barts Heath NHS Trust |
| Raghavendran Kulasegaran-Shylini | Barts Heath NHS Trust |
| Claire E. Broad | Barts Heath NHS Trust |
| Dola Owoyemi | Barts Heath NHS Trust |
| David Harrington | Barts Heath NHS Trust |
| Clare Coffey | Barts Heath NHS Trust |
| Martina Cummins | Barts Heath NHS Trust |
| Anna Riddell | Barts Heath NHS Trust |
| Tyrra D'Souza | Barts Heath NHS Trust |

**Glasgow site**

| Name of individual | Employing Institution |
|---|---|
| Guy Mollett | MRC-University of Glasgow Centre for Virus Research |
| **Emma Thomson** | **MRC-University of Glasgow Centre for Virus Research and NHS Greater Glasgow and Clyde** |
| Christine Peters | NHS Greater Glasgow and Clyde |
| Aleks Marek | NHS Greater Glasgow and Clyde |
| Rory Gunson | NHS Greater Glasgow and Clyde |
| Emily Goldstein | NHS Greater Glasgow and Clyde |
| Emilie Shepherd | NHS Greater Glasgow and Clyde |
| James Shepherd | MRC-University of Glasgow Centre for Virus Research |
| David Robertson | MRC-University of Glasgow Centre for Virus Research |
| Katherine Smollett | MRC-University of Glasgow Centre for Virus Research |
| Ana da Silva Filipe | MRC-University of Glasgow Centre for Virus Research |
| Alice Broos | MRC-University of Glasgow Centre for Virus Research |
| Stephen Carmichael | MRC-University of Glasgow Centre for Virus Research |
| Nicholas Suarez | MRC-University of Glasgow Centre for Virus Research |
| Chris Davis | MRC-University of Glasgow Centre for Virus Research |

| | |
|---|---|
| Sreenu Vattipally | MRC-University of Glasgow Centre for Virus Research |
| Joseph Hughes | MRC-University of Glasgow Centre for Virus Research |
| Ioulia Tsatsani | MRC-University of Glasgow Centre for Virus Research |
| Jacqueline McTaggart | NHS Greater Glasgow and Clyde |
| Stephanie McEnhill | NHS Greater Glasgow and Clyde |

**Guy's and St Thomas' site**

| Name of individual | Employing Institution |
|---|---|
| Adela Medina | Viapath |
| Themoula Charalampous | KCL |
| Bindi Patel | GSTT NHS Trust |
| Flavia Flaviani | GSTT NHS Trust |
| Jörg Saßmannshausen | GSTT NHS Trust |
| May Rabuya | GSTT NHS Trust |
| Sulekha Gurung | GSTT NHS Trust |
| Anu Augustine | GSTT NHS Trust |
| Rahul Batra | GSTT NHS Trust |
| Luke Snell | GSTT NHS Trust |
| **Gaia Nebbia** | **GSTT NHS Trust** |

**Imperial site**

| Name of individual | Employing Institution |
|---|---|
| **Alison Holmes** | **Imperial Healthcare NHS Trust** |
| Sid Mookerjee | Imperial Healthcare NHS Trust |
| James Price | Imperial Healthcare NHS Trust |
| Paul Randell | Imperial Healthcare NHS Trust |
| Krystal Johnson | Imperial Healthcare NHS Trust |
| Thilipan Thaventhiran | Imperial Healthcare NHS Trust |
| Damien Mine | Imperial Healthcare NHS Trust |
| Sophie Hunter | Imperial Healthcare NHS Trust |
| Isa Ahmad | Imperial Healthcare NHS Trust |
| Anitha Ramanathan | Imperial Healthcare NHS Trust |

**Liverpool site**

| Name of individual | Employing Institution |
|---|---|
| **Anu Chawla** | **Liverpool NHS Foundation Trust** |
| Alistair Darby | University of Liverpool |
| Sam Haldenby | University of Liverpool |

| | |
|---|---|
| Mark Whitehead | University of Liverpool |
| Claudia Wierzbicki | University of Liverpool |
| Hermione Webster | University of Liverpool |
| Amy Colleran | University of Liverpool |
| Flora Todd | University of Liverpool |
| Hannah Trivett | University of Liverpool |
| Anita Lucaci | University of Liverpool |
| Becky Taylor | Liverpool NHS Foundation Trust |
| Keith Morris | Liverpool NHS Foundation Trust |
| Charles Numbere | Liverpool NHS Foundation Trust |
| Mark Hopkins | Liverpool NHS Foundation Trust |
| Jenifer Mason | Liverpool NHS Foundation Trust |
| Alexandra Bailey | Liverpool NHS Foundation Trust |
| Debbie Lankstead | Liverpool NHS Foundation Trust |
| Damian Burns | Liverpool NHS Foundation Trust |

**Manchester site**

| Name of individual | Employing Institution |
|---|---|
| **Nicholas Machin** | **PHE and Manchester University NHS Foundation Trust** |
| Shazaad Ahmad | Manchester University NHS Foundation Trust |
| Julie Cawthorne | Manchester University NHS Foundation Trust |
| Ryan George | Manchester University NHS Foundation Trust |
| James Montgomery | Manchester University NHS Foundation Trust |
| Deborah McKew | Manchester University NHS Foundation Trust |
| Osama Abdul-Wahab | PHE |
| Thomas El-Basha | PHE |
| James Barnes | PHE |
| Ian Venables | Manchester University NHS Foundation Trust |
| Katharine Wylie | Manchester University NHS Foundation Trust |
| Karen Talbot | Manchester University NHS Foundation Trust |

**Newcastle site**

| Name of individual | Employing Institution |
|---|---|
| **Yusri Taha** | **Newcastle NHS Trust** |
| Angela Cobb | Newcastle NHS Trust |

| | |
|---|---|
| Michelle Ramsay | Newcastle NHS Trust |
| Maria Leader | Newcastle NHS Trust |
| Shirelle Burton-Fanning | Newcastle NHS Trust |
| Julie Samuel | Newcastle NHS Trust |
| Milo Cullinan | Newcastle NHS Trust |
| Sarah Francis | Newcastle NHS Trust |
| Lydia Taylor | Newcastle NHS Trust |
| Darren Smith | Northumbria University |
| Matthew Bashton | Northumbria University |
| Matthew Crown | Northumbria University |
| Andrew Nelson | Northumbria University |
| Clare McCann | Northumbria University |
| Gregory Young | Northumbria University |
| Will Stanley | Northumbria University |
| Zack Richards | Northumbria University |
| Rui Dos Santos | Northumbria University |

**Nottingham site**

| Name of individual | Employing Institution |
|---|---|
| **Nikunj Mahida** | **Nottingham NHS Trust** |
| William Irving | Nottingham NHS Trust |
| Matthew Loose | University of Nottingham |
| Patrick McClure | University of Nottingham |
| Mitch Clarke | Nottingham NHS Trust |
| Elaine Baxter | Nottingham NHS Trust |
| Carl Yates | Nottingham NHS Trust |
| Irfan Aslam | Nottingham NHS Trust |
| Vicki Fleming | Nottingham NHS Trust |
| Michelle Lister | Nottingham NHS Trust |
| Johnny Debebe | University of Nottingham |
| Nadine Holmes | University of Nottingham |
| Christopher Moore | University of Nottingham |
| Matt Carlile | University of Nottingham |

**Royal Free site**

| Name of individual | Employing Institution |
|---|---|

| | |
|---|---|
| **Tabitha Mahungu** | **Royal Free London NHS Trust** |
| Sophie Weller | Royal Free London NHS Trust |
| Tanzina Haque | Royal Free London NHS Trust |
| Jennifer Hart | Royal Free London NHS Trust |
| Dianne Irish-Tavares | Royal Free London NHS Trust |
| Eric Witele | Royal Free London NHS Trust |
| Mia De Mesa | Royal Free London NHS Trust |
| Vicky Pang | Royal Free London NHS Trust |
| Jelena Heaphy | Royal Free London NHS Trust |
| Wendy Chatterton | Health Services Laboratory |
| Monika Pusok | Health Services Laboratory |

**Sandwell site**

| Name of individual | Employing Institution |
|---|---|
| **Dr Tranprit Saluja** | **Sandwell & West Birmingham Hospitals NHS Trust** |
| Zahira Maqsood | Sandwell NHS Trust |
| Angie Williams | Sandwell NHS Trust |
| Debbie Devonport | Sandwell NHS Trust |
| Lucy Palinkas | Sandwell NHS Trust |
| Diane Thomlinson | Sandwell NHS Trust |
| Julie Booth | Sandwell NHS Trust |
| Ashok Dadrah | Sandwell NHS Trust |
| Amanda Symonds | Sandwell NHS Trust |
| Cassandra Craig | Sandwell NHS Trust |
| Dr Abhinav Kumar | Sandwell NHS Trust |

**Sheffield site**

| Name of individual | Employing Institution |
|---|---|
| **Thushan de Silva** | **University of Sheffield** |
| Matthew D Parker | University of Sheffield |
| Peijun Zhang | University of Sheffield |
| Max Whiteley | University of Sheffield |
| Benjamin B Lindsey | University of Sheffield |
| Paige Wolverson | University of Sheffield |
| Benjamin H Foulkes | University of Sheffield |
| Luke Green | University of Sheffield |
| Marta Gallis Ramalho | University of Sheffield |
| Stavroula F Louka | University of Sheffield |
| Adrienn Angyal | University of Sheffield |
| Nikki Smith | University of Sheffield |

| | |
|---|---|
| David G Partridge | Sheffield NHS Trust |
| Cariad Evans | Sheffield NHS Trust |
| Mohammad Raza | Sheffield NHS Trust |
| Hayley Colton | Sheffield NHS Trust |
| Rebecca Gregory | Sheffield NHS Trust |
| Phillip Ravencroft | Sheffield NHS Trust |
| Katie Johnson | Sheffield NHS Trust |
| Sharon Hsu | University of Sheffield |
| Alexander J Keeley | Sheffield NHS Trust |
| Alison Cope | Sheffield NHS Trust |
| Amy State | Sheffield NHS Trust |
| Nasar Ali | Sheffield NHS Trust |
| Rasha Raghei | Sheffield NHS Trust |
| Joe Heffer | Sheffield NHS Trust |
| Stella Christou | University of Sheffield |
| Samantha E Hansford | University of Sheffield |
| Hailey R Hornsby | University of Sheffield |
| Phil Wade | Sheffield NHS Trust |
| Kay Cawthron | Sheffield NHS Trust |
| Maqsood Khan | Sheffield NHS Trust |
| Amber Ford | Sheffield NHS Trust |
| Imogen Wilson | Sheffield NHS Trust |
| Kate Harrington | Sheffield NHS Trust |
| Nic Tinker | Sheffield NHS Trust |
| Sally Nyinza | Sheffield NHS Trust |

**Southampton site**

| Name of individual | Employing Institution |
|---|---|
| **Kordo Saeed** | University Hospital Southampton NHS Foundation Trust |
| Jacqui Prieto | University Hospital Southampton NHS Foundation Trust |
| Adhyana Mahanama | University Hospital Southampton NHS Foundation Trust |
| Buddhini Samaraweera | University Hospital Southampton NHS Foundation Trust |
| Siona Silviera | University Hospital Southampton NHS Foundation Trust |
| Emanuela Pelosi | University Hospital Southampton NHS Foundation Trust |
| Eleri Wilson-Davies | University Hospital Southampton NHS Foundation Trust |
| Sarah Jeremiah | University Hospital Southampton NHS Foundation Trust |
| Helen Wheeler | University Hospital Southampton NHS Foundation Trust |

| | |
|---|---|
| Matthew Harvey | University Hospital Southampton NHS Foundation Trust |
| Thea Sass | University Hospital Southampton NHS Foundation Trust |
| Helen Umpleby | University Hospital Southampton NHS Foundation Trust |
| Stephen Aplin | University Hospital Southampton NHS Foundation Trust |
| Samuel Robson | University of Portsmouth |
| Sharon Glaysher | Portsmouth Hospital NHS Trust |
| Scott Elliott | Portsmouth Hospital NHS Trust |
| Kate Cook | University of Portsmouth |
| Christopher Fearn | University of Portsmouth |
| Salman Goudarzi | University of Portsmouth |
| Katie Loveson | University of Portsmouth |
| Angela Beckett | University of Portsmouth |

**St George's site**

| Name of individual | Employing Institution |
|---|---|
| Kenneth Laing | St Georges, UoL |
| Irene Monahan | St Georges, UoL |
| Adam Witney | St Georges, UoL |
| Joshua Taylor | St Georges NHS Trust |
| NgeeKeong Tan | St Georges NHS Trust |
| **Cassie Pope** | **St Georges NHS Trust and St Georges, UoL** |
| Claudia Cardosa Pereira | St Georges NHS Trust |
| Vaz Malik | St Georges, UoL |

**UCLH site**

| Name of individual | Employing Institution |
|---|---|
| **Gee Yen Shin** | **UCLH NHS Trust** |
| Eleni Nastouli | UCLH NHS Trust |
| Catherine Houlihan | UCLH NHS Trust |
| Judith Heaney | UCLH NHS Trust |
| Matthew Byott | UCLH NHS Trust |
| Dan Frampton | UCL / UCLH |
| Gema Martinez-Garcia | UCLH NHS Trust |
| Leila Hail | UCLH NHS Trust |
| Ndifreke Atang | UCLH NHS Trust |
| Helen Francis | UCLH NHS Trust |
| Milica Rajkov | UCLH NHS Trust |

**UCL Genomics**

| Name of individual | Employing Institution |
|---|---|
| **Judith Breuer** | **UCL** |
| Rachel Williams | UCL |
| Sunando Roy | UCL |
| Charlotte Williams | UCL |
| Nadua Bayzid | UCL |
| Marius Cotic | UCL |

**UCL Comprehensive Clinical Trials Unit**

| Name of individual | Employing Institution |
|---|---|
| James Blackstone | UCL |
| Leanne Hockey | UCL |
| Alyson MacNeil | UCL |
| Rachel McComish | UCL |
| Monica Panca | UCL |
| Georgia Marley | UCL |

**UCL Institute for Global Health**

| Name of individual | Employing Institution |
|---|---|
| Andrew Copas | UCL |
| Oliver Stirrup | UCL |
| Fiona Mapp | UCL |

**UCL Research IT Services**

| Name of individual | Employing Institution |
|---|---|
| Asif Tamuri | UCL |
| Stefan Piatek | UCL |

**University of Strathclyde**

| Name of individual | Employing Institution |
|---|---|
| Paul Flowers | UoS |

**Francis Crick Institute**

| Name of individual | Employing Institution |
|---|---|
| Marg Crawford | Francis Crick Institute |
| Laura Cubitt | Francis Crick Institute |
| Deborah J Jackson | Francis Crick Institute |
| Jimena Perez-Lloret | Francis Crick Institute |

| | |
|---|---|
| Sophie Ward | Francis Crick Institute |
| Makis Fidanis | Francis Crick Institute |
| Aaron Sait | Francis Crick Institute |
| Robert Goldstone | Francis Crick Institute |
| Harshil Patel | Francis Crick Institute |
| Chelsea Sawyer | Francis Crick Institute |
| Aengus Stewart | Francis Crick Institute |
| Steve Gamblin | Francis Crick Institute |
| Charles Swanton | Francis Crick Institute |
| Jerome Nicod | Francis Crick Institute |
